# RNA-sequencing first approach generates new diagnostic candidates in Mendelian disorders

**DOI:** 10.1101/2023.07.05.23292254

**Authors:** Carolina Jaramillo Oquendo, Htoo A Wai, Will Rich, David J. Bunyan, N. Simon Thomas, David Hunt, Jenny Lord, Andrew G L Douglas, Diana Baralle

## Abstract

**Background:** RNA-sequencing is increasingly being used as a complementary tool to DNA sequencing in diagnostics where DNA analysis has been uninformative. RNA-sequencing allows us to identify alternative splicing and aberrant gene expression allowing for improved interpretation of variants of unknown significance (VUS). Additionally, RNA-sequencing provides the opportunity not only to look at the splicing effects of known VUSs but also to scan the transcriptome for abnormal splicing events and expression abnormalities in other relevant genes that may be the cause of a patient’s phenotype.

**Methods:** Using RNA from patient blood, we have systematically assessed transcriptomic profiles of 87 patients with suspected Mendelian disorders, 38% of which did not have a candidate sequence variant. Cases with VUSs and known events were assessed first followed by assessment of cases with no VUS. Each VUS was visually inspected using the Integrative Genomics Viewer (IGV) to search for splicing abnormalities. Once aberrant splicing was identified in cases with VUS, multiple open-source alternative splicing tools (MAJIQ, rMATS-turbo, FRASER2 and LeafCutterMD) were used to investigate if they would identify what was observed in IGV. Expression outliers were detected using OUTRIDER. To find diagnoses in cases without a VUS or gene of interest, two separate strategies were used. The first was a genotype to phenotype approach using variant calls obtained from the RNA-sequencing and overlapping those calls with results from splicing tools. The second strategy involved using phenotype information available to filter results from splicing tools.

**Results:** Using RNA-sequencing only, we were able to assess 71% of VUSs and detect aberrant splicing in 14/48 patients with a VUS. Furthermore, we identified four new diagnoses by detecting novel aberrant splicing events in patients with no candidate sequence variants from prior genomic DNA testing (n=33) or those in which the candidate VUS did not affect splicing (n=23) and identified one additional diagnosis through detection of skewed X-inactivation.

**Conclusion:** We demonstrate the identification of novel diagnoses using an RNA-sequencing first approach in patients without candidate VUSs. Furthermore, we demonstrate the utility of blood-based RNA analysis in improving diagnostic yields and highlight optimal approaches for such analysis.

## Background

With the advancement of next generation sequencing, vast amounts of DNA sequencing data are continually generated to aid in the diagnosis and treatment of rare diseases. However, our ability to interpret genomic data has not grown at the same rate. The diagnostic yield of whole exome and genome sequencing alone remains relatively low leaving scope for diagnostic rates to be improved(1–4). Within the Genomics England 100,000 Genomes Project for example, the average diagnostic yield using whole genome sequencing is around 25%(3). RNA sequencing (RNA-seq) is now being used as a complementary tool to DNA sequencing for diagnostic genetic testing in rare disease where DNA analysis alone has failed to identify a clear diagnosis(5–15). While some studies have focused on specific disorder types, such as mitochondrial disease(6,12), muscle disorders(5), and neurodevelopmental disorders(13), others have looked at heterogeneous disease populations(7,11,14–16). Unlike DNA sequencing, high throughput RNA-seq is both a qualitative and quantitative approach which allows identification of aberrant splicing (AS), aberrant gene expression, and mono allelic expression, allowing improved interpretation of variants of unknown significance (VUSs). An important benefit of the transcriptomic approach as compared to targeted reverse transcription PCR (RT-PCR) is that it is agnostic to the resulting abnormally spliced transcript, whereas RT-PCR must rely on targeted primer designs that are intrinsically limited by factors such as known gene annotations, PCR amplicon lengths and expected aberrant splicing event. RNA-seq therefore provides the opportunity not only to look at the splicing effects of known VUSs but also to scan the transcriptome for abnormal splicing events and expression abnormalities in other relevant genes that may be the cause of a patient’s phenotype. This in turn allows the identification of molecular diagnoses in patients in which standard genomic DNA testing has not identified any candidate.

In this study, we have used RNA-seq to a) investigate the feasibility of generating new diagnostic candidates in a subset of patients with no previously reported candidate VUSs in clinically relevant genes and; b) assess the use of blood as the tissue of choice in the implementation of an RNA-seq clinical pipeline to improve diagnostic yield of patients with rare diseases. To understand limitations of available tools we first examined the splicing effects of clinically relevant VUSs in a heterogeneous cohort of patients with suspected Mendelian disorders.

## Material and Methods

### Patient recruitment

Participants were enrolled into the University of Southampton’s Splicing and Disease study with appropriate ethical approval (REC 11/SC/0269, IRAS 49685, ERGO 23056). This cohort of individuals comprises a combination of rare disease patients assessed by UK clinical genetics services in whom a candidate VUS may or may not have been identified through conventional DNA-based testing (n=87). Within this cohort, 48 individuals had pre-existing candidate VUSs (n=51 variants) that had been previously clinically reported within genes of potential clinical relevance, six had previously confirmed known genetic diagnoses (4 array deletions, 1 PURA duplication and 1 PURA deletion) and 33 had unknown molecular diagnoses with no previously reported candidate VUSs in clinically relevant genes. Individuals without molecular diagnosis had a phenotype where a genetic cause was suspected. 18 of the 51 VUSs have been previously assessed by RT-PCR(10).

### Sample collection and RNA extraction

RNA was extracted from 87 blood samples collected in PAXgene blood RNA tubes using the PAXgene blood RNA Kit (PreAnalytiX, Switzerland). Quantification was performed by NanoDrop spectrophotometer and Qubit fluorometer (Thermo, MA) and RNA integrity number assessed using an Agilent 2100 Bioanalyzer (Agilent, CA).

### RNA sequencing

RNA samples were sequenced via Novogene (Hong Kong) in four separate batches (comprising 7, 16, 33 and 31 samples) using a total RNA-seq approach employing the NEBNext rRNA Depletion Kit and the NEBNext Ultra Directional RNA Library Prep Kit (New England Biolabs, MA). Samples in batches 1, 2 and 4 also had NEBNext Globin Depletion Kit applied, whereas those in batch 3 did not. The library was checked with Qubit and real-time PCR for quantification and bioanalyzer for size distribution detection. On average 76 million 150 base-pair paired-end reads were generated for each sample on a HiSeq 2000 instrument (Illumina, CA). FASTQ files underwent initial quality control filtering and adapter sequence removal by Novogene. Filtering included removal of reads containing N>10% (N: bases that cannot be determined) and reads with over 50% of low quality (Qscore ≤ 5) bases. Subsequent alignment was performed to the human genome reference (GRCh38) with annotations from GENCODE(17) release 38 using STAR aligner(18) v2.6.1c with optimised parameters via the University of Southampton’s IRIDIS5 high-performance computing clusters. Scripts can be found in GitHub [https://github.com/carojoquendo/RNA_splicing_and_disease].

### Assessment of aberrant splicing in cases with VUS and known molecular diagnosis

Due to the nature of the cohort, assessment of aberrant splicing and expression was done in stages. Cases with VUSs and known events were assessed first followed by assessment of cases with no VUS (**Figure 1**).

**Figure 1.**
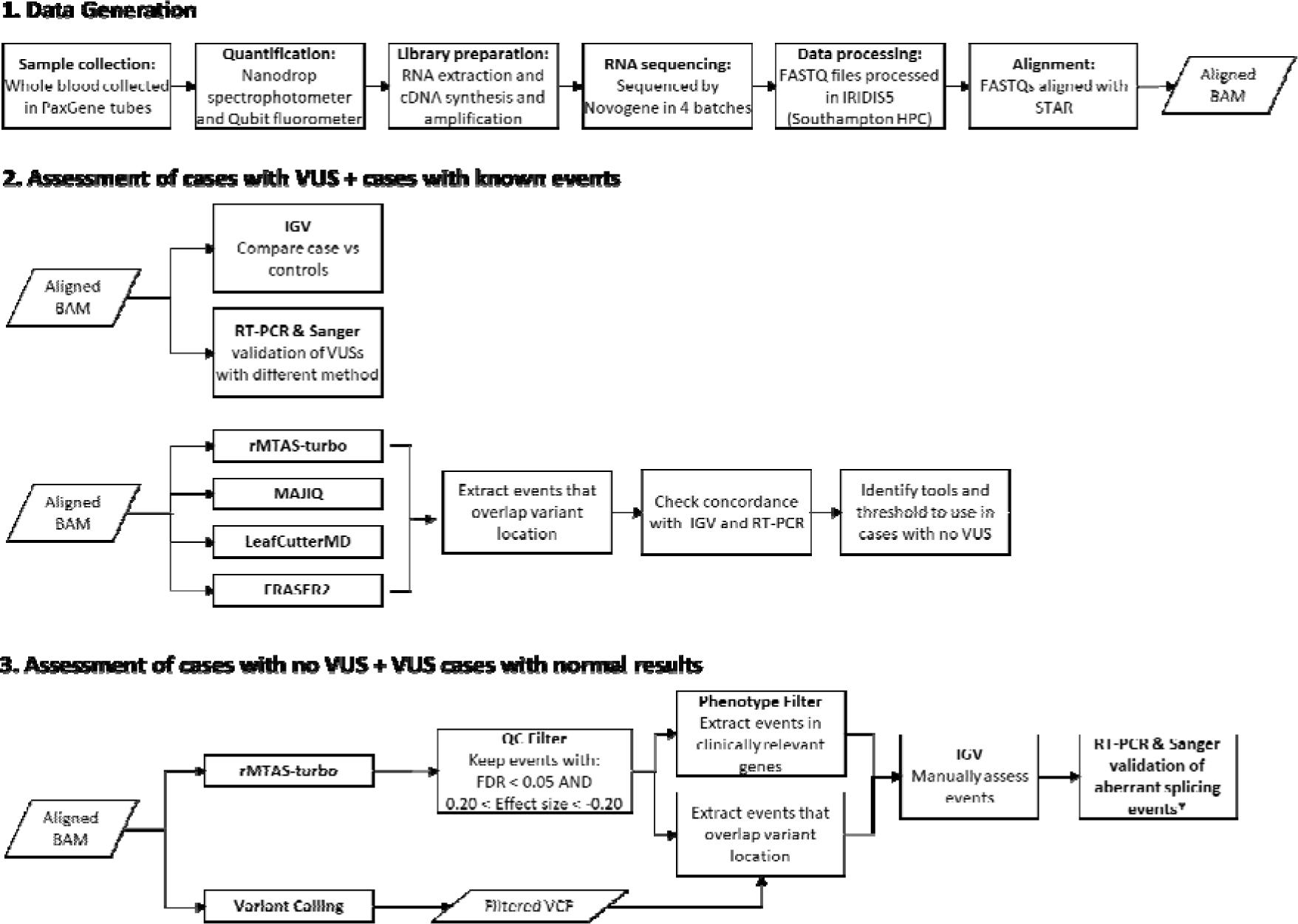
Overview of methods. **1.** Data generation. This step was generally the same for all samples (n=87). The only difference came in the library preparation stage where samples in batches 1, 2 and 4 also had globin depletion, whereas samples in batch 3 did not. **2.** Cases with a VUS were assessed first. Each variant of unknown significance (VUS) was visually inspected to search for splicing abnormalities in the Integrative Genomics Viewer (IGV). RT-PCR and Sanger sequencing were carried out in parallel for additional validation. **3.** Cases without a clinically relevant candidate variant were investigated last. This also included cases for which the original candidate VUS had not been found to alter splicing. Filtering strategy was determined based on observations across cases with a VUS. Results were visualised in IGV, and new diagnostic candidates were validated with RT-PCR and Sanger sequencing.

To determine the functional consequence at a transcript level for each VUS, RNA-seq data was loaded into the Integrative Genomics Viewer (IGV)(19) and each variant was visually inspected to search for splicing abnormalities. If there were no splicing abnormalities in the exons and introns flanking the variant, it was determined that there were no splicing abnormalities resulting from the variant. Splicing abnormalities were classed as: exon skipping, inclusion of cryptic exon, intron retention, alternative 5ill (donor) splice site and alternative 3ill (acceptor) splice site. The command-line tool ggsashimi(20) was used to create final sashimi plots to visualise junctions. RT-PCR was carried out in parallel to assess VUSs where possible. In some cases, RT-PCR was not carried out due to technical limitations (additional details can be found in the supplemental table 1).

Once aberrant splicing events had been ascertained in cases that had a VUS or known molecular diagnosis, we used this data to identify open-source tools best placed to identify potential aberrant splicing in cases for which there was no candidate variant. FRASER2(21,22), rMATS-turbo v4.1.2(23), MAJIQ v2.4(24) and LeafCutterMD v0.2.9(25) were used to detect aberrant splicing across all samples. The tools chosen are some of the most commonly used for splicing analyses, where rMATS-turbo and MAJIQ are events-based methods while LeafCutterMD and FRASER2 are outlier approaches. We decided to use tools with two different underlying methodologies as there is still no gold standard for identifying splicing events in this type of cohort. For all tools except FRASER2, each sample was compared against other samples within the same batch with the exception of samples in batch 1 and 2 which were combined to increase power. rMATS-turbo was run with additional parameters--novelSS to enable detection of novel splice sites, as well as--allow-clipping to allow alignments with soft or hard clipping to be used. MAJIQ modules build and DeltaPSI were run with default parameters using the GENCODE v38 annotation gff3 files.

The DeltaPSI results were then input into the MAJIQ voila module which provides a tab-delimited text file to allow parsing of the MAJIQ result and filters out local splice variations (LSVs) with no junctions predicted to change over a certain value. Default parameters for the voila module were used. LeafCutterMD and FRASER2 were also run with default parameters and results were annotated with gene symbols to extract genes and loci of interest for each sample. Once samples had been run through all the tools, events within the gene of interest were extracted to determine if the tools had been able to pick up what had been seen in IGV. This allowed us to check concordance between the tools and identify thresholds that could be used later when looking for events in cases without a VUS.

### Assessment of aberrant splicing in patients without candidate VUSs

After cases with VUSs and known events were assessed, we investigated those without a clinically relevant candidate variant. This also included cases for which the original candidate VUS had not been found to alter splicing. The aligned BAM files were run through rMATS-turbo as well as through the GATK’s Best Practices workflow for RNA-seq short variant discovery (https://gatk.broadinstitute.org/hc/en-us/articles/360035531192-RNAseq-short-variant-discovery-SNPs-Indels-) to identify variants in the RNA-seq.

#### Variant calling

First duplicate reads were marked by Picard’s version 2.18.14 (http://broadinstitute.github.io/picard) MarkDuplicates function, followed by reformatting of the BAM files for HaplotypeCaller with GATK’s (version 4.2.2)(26) SplitNCigarReads and Picard’s AddOrReplaceReadGroups. The next step was Base Quality Recalibration, consisting of two tools: GATK’s BaseRecalibrator and ApplyBQSR. Lastly GATK’s HaplotypeCaller was used to call variants and write to VCF files.

To reduce spurious calls, VCF files were run through GATK’s VariantFiltration tool, keeping calls with a minimum quality score of 50. bcftools(27) was used to further filter variants excluding any variants with a) less than eight reads covering the locus; b) calls with genotype quality lower than 16; c) calls with strand bias (FS metric) greater than 30; and d) variants with a quality normalised by depth of at least two(28). After filtering, Ensembl’s VEP(29) (version 103) was used to annotate the variants with additional information including but not limited to nearest gene, variant consequence (e.g. missense, splice_region) and minor allele frequency (MAF). The SpliceAI VEP plugin was used to produce a score per variant (delta score) based on the likelihood of the variant impacting splicing. SpliceAI scores range from 0-1 with scores closer to 1 being more likely to affect splicing. VCF files were further filtered to keep variants that met all of the following conditions: a) population frequency less than 0.005; b) variants with a SpliceAI score ≥ 0.2; c) variants found in protein coding genes; and d) single nucleotide variants. Indels were not included as the majority had very poor quality and inclusion of indels introduced a significant number of false positives.

#### Filtering strategies

To find diagnoses in cases without a VUS or gene of interest, two separate strategies were used. The first was a genotype to phenotype approach. Using the annotated and filtered VCF files a BED file was created with variant location, adding 25 base pairs up and downstream of the variant [chromosome start(−25bp) end(+25bp) gene]. rMATS-turbo results were also converted into BED format. After sorting the BED files, the variant BED was overlapped with the rMATS-turbo results BED using bedtools (v2.30)(30) intersect keeping only overlapping features. Each genomic location that was found to overlap a variant and an alternative splicing event identified by rMATS-turbo was then inspected in IGV as previously described.

The second strategy involved using phenotype information available to filter results from splicing tools. To do this, appropriate panels from the UK Genomic Medicine Service (GMS) PanelApp(31) resource were applied to the splicing tools results and each AS event was also inspected in IGV.

### Minimum number of sequencing reads & splice effect predictions

The MRSD web portal (https://mcgm-mrsd.github.io/) was used to predict the minimum number of sequencing reads required from RNA-seq experiments to confidently determine aberrant splicing events for a gene of interest(32). Default values for confidence level (95%) and splice junction proportion (75%) were used. For coverage a minimum of five reads were used (n=5). The online SpliceAI server (https://spliceailookup.broadinstitute.org/) was used to predict splicing effect of all variants.

### RT-PCR and Sanger sequencing

Primers for the variant of interest were designed to span at least three exons and where possible, up to seven exons. The cDNA was synthesised using High-Capacity cDNA Reverse Transcription Kit (Thermo Fisher Scientific, USA). PCR was performed using GoTaq G2 DNA polymerase kit (Promega, USA). The PCR products were analysed in a 1% agarose gel prepared with Nancy-520 DNA gel stain (Sigma, USA). Subsequently, the PCR products were purified using the GeneJET PCR Purification Kit (Thermo Fisher Scientific, USA) and bidirectional Sanger sequencing was carried out by SourceBioscience. PCR experiments were repeated for reproducibility.

### Expression outlier detection

Salmon v1.6.0(36) was used to quantify gene and transcript counts in mapping-based mode. Transcriptome indices for Salmon were generated using the GRCh38 genome and transcriptome reference from GENCODE release 38 (https://combine-lab.github.io/alevin-tutorial/2019/selective-alignment/). The R v 4.1.1(37) package tximport v1.22.0(38) was used to collate and import raw read counts from all samples to be used as input into OUTRIDER v1.12.0(39). The OUTRIDER function filterExpression was used to remove genes that had low Fragments Per Kilobase of transcript per Million mapped reads (FPKM) expression values followed by the OUTRIDER function which ran the full OUTRIDER pipeline.

## Results

### Summary of RNA-sequencing data outputs

The mean number of sequencing reads per sample was 76.6 million (61.3-130.2 million) and on average 80% of reads were uniquely mapping (**Figure S1 A & B**). Mean number of splicing junctions identified across samples (**Figure S1 C**) was 398,718 (303,637–621,161). Spearman’s rank correlation between observed median TPM values and median TPM values found in the Genotype-Tissue Expression (GTEx) portal(40) was 0.79 with a p-value < 0.001 (**Figure S1 D**). When considering disease genes from the Online Mendelian Inheritance in Man (OMIM) database(41) and the UK Genomic Medicine Service’s PanelApp resource, 67% (n=11,128) and 75% (n=2,721) of genes were expressed in blood respectively (TPM >1 in at least 4 samples).

As mentioned previously, globin depletion was not applied to one of the batches (batch 3). Analysis of the transcriptomic profiles showed this difference in targeting methodology as samples within batch 3 clustered together in principal component analysis as well as hierarchical clustering (**Figure S1 E & F**). Furthermore, median TPM values for the most abundant haemoglobin genes were in line with values reported in GTEx, which also did not utilise globin depletion. To avoid bias due to differences in sequencing methodology, samples were run in separate batches through the splicing tools. TPM values across genes which had VUSs within our cohort were also assessed (**Figure S2**), which showed that gene coverage in genes of interest was not negatively affected by the lack of globin depletion and in fact in some cases, the TPM values were higher in batch 3. A possible explanation for the slight increase of reads in the non-depleted batch is that the globin depletion step could also be reducing the reads for some non-haemoglobin genes as well(42). A comparison of the whole transcriptome between batch 3 and the other three batches (**Figure S3**) demonstrated that globin depletion increased coverage of lower expressed genes (median TPM 0-1), but in general the lack of globin depletion does not seem to have a large effect on the transcriptomic profiles.

### Splicing analysis in patients with a candidate VUS

We began by looking at the 48 cases which had a VUS to guide our analysis of those for which we had no candidate variant. This entailed investigation of 51 VUSs across 36 different genes, as some cases had more than one VUS. Using default parameters, the MRSD tool predicted that we would only be able to assess 47% (n=17) of the 36 genes in blood based on our mean number of sequencing reads. However, we found this tool to be overly conservative as we were able to assess 69% of genes (n=25) using RNA-seq alone. Using the GTEx dataset as a reference(43), median TPM values across the 25 genes ranged from 0.89 - 73.24 with a mean and median of 19.43 and 11.16 respectively. The experimental median TPM values in our sequencing data ranged from 2.142 – 75.896 with a mean and median of 15.718 and 7.637 respectively.

Visual inspection of the RNA-seq BAM files in IGV allowed the detection of aberrant splicing in 14 cases with candidate VUSs (**Table 1**). Of these splice-altering VUSs, 13/14 were predicted to affect splicing according to SpliceAI (Δ score ≥ 0.2) and all 14 were validated via RT-PCR (RT-PCR results for 6 of the 14 variants have been previously reported(44)). The gene with the lowest median TPM value in GTEx for which we were able to detect aberrant splicing was KAT6B with a GTEx TPM of 0.890 and an experimental median TPM value of 9.79. Out of the 38 VUSs where aberrant splicing events were not detected, 15 variants could not be assessed as a result of low gene expression in blood (<10 reads covering locus or normal junctions not observed in sample and controls). For genes where RNA seq was uninformative, median TPM values according to GTEx ranged from 0.00 to 4.91 with a mean and median of 0.49 and 0.08 respectively. RT-PCR was able to validate aberrant splicing in four additional cases with variants in TERT, PRG4, and TAOK1. Details of all assessed variants can be found in **Table S1**. Cases (n=23) which showed no aberrant splicing linked to a VUS were subsequently analysed as unknown cases.

**Table 1.**
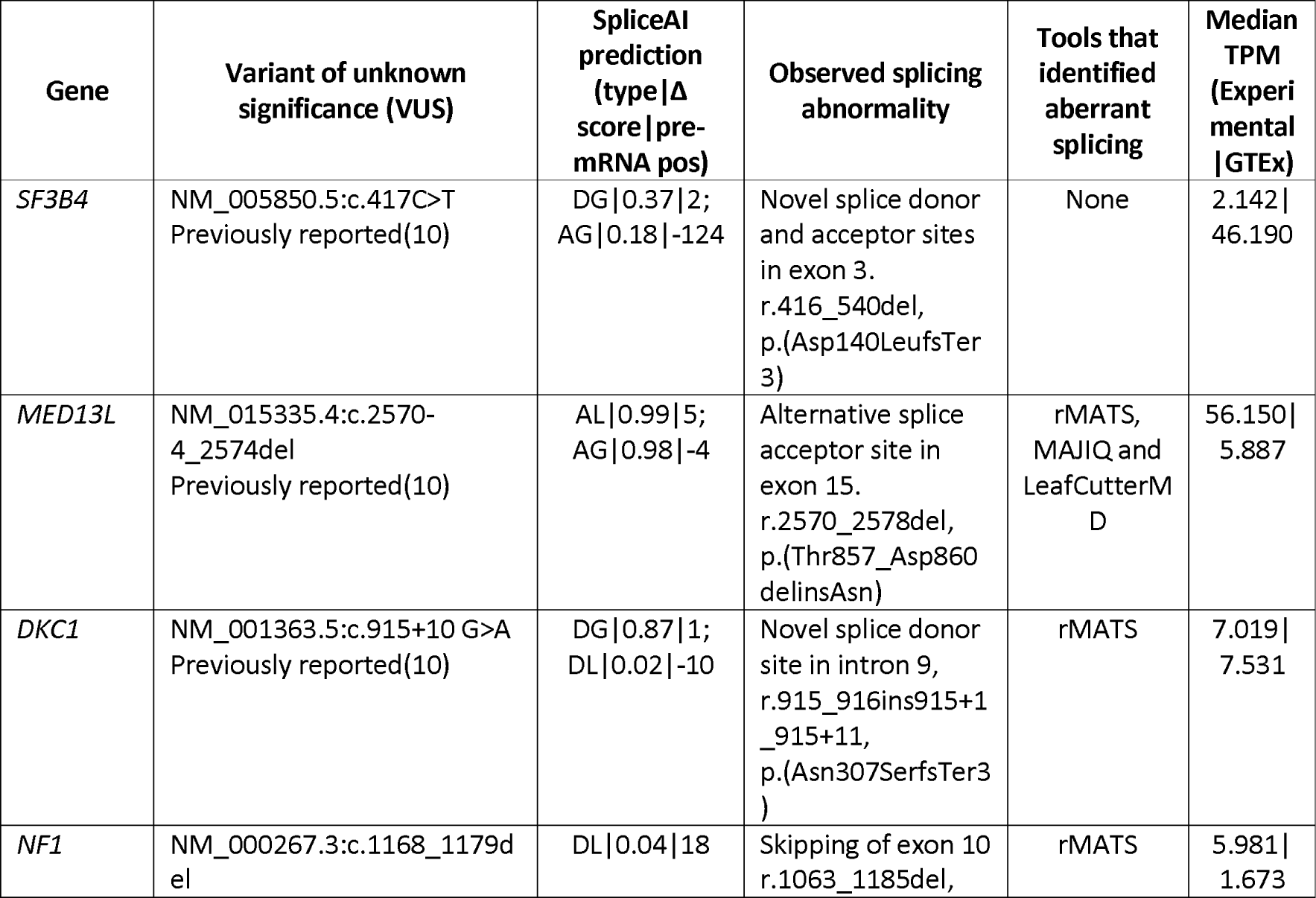
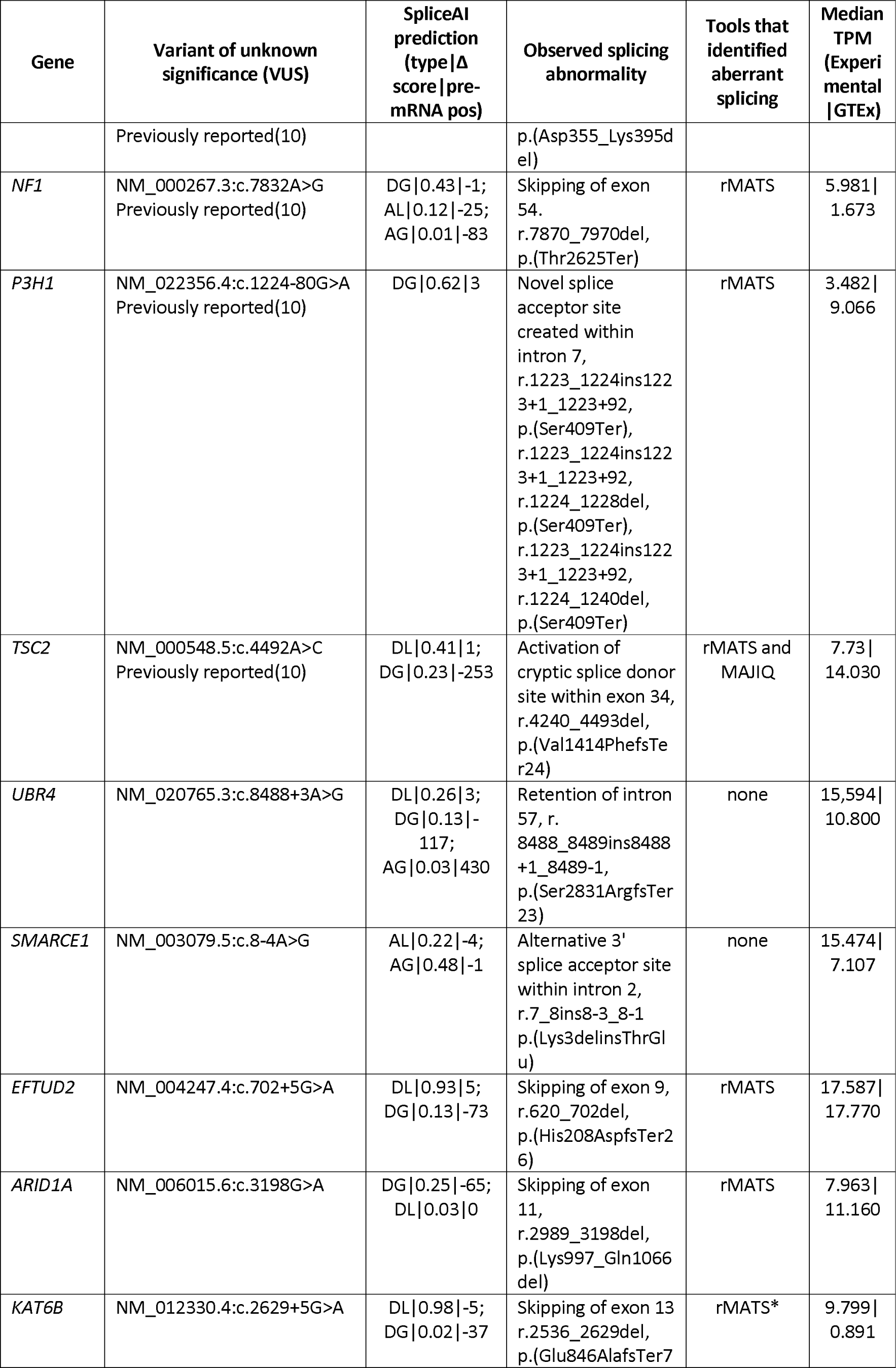
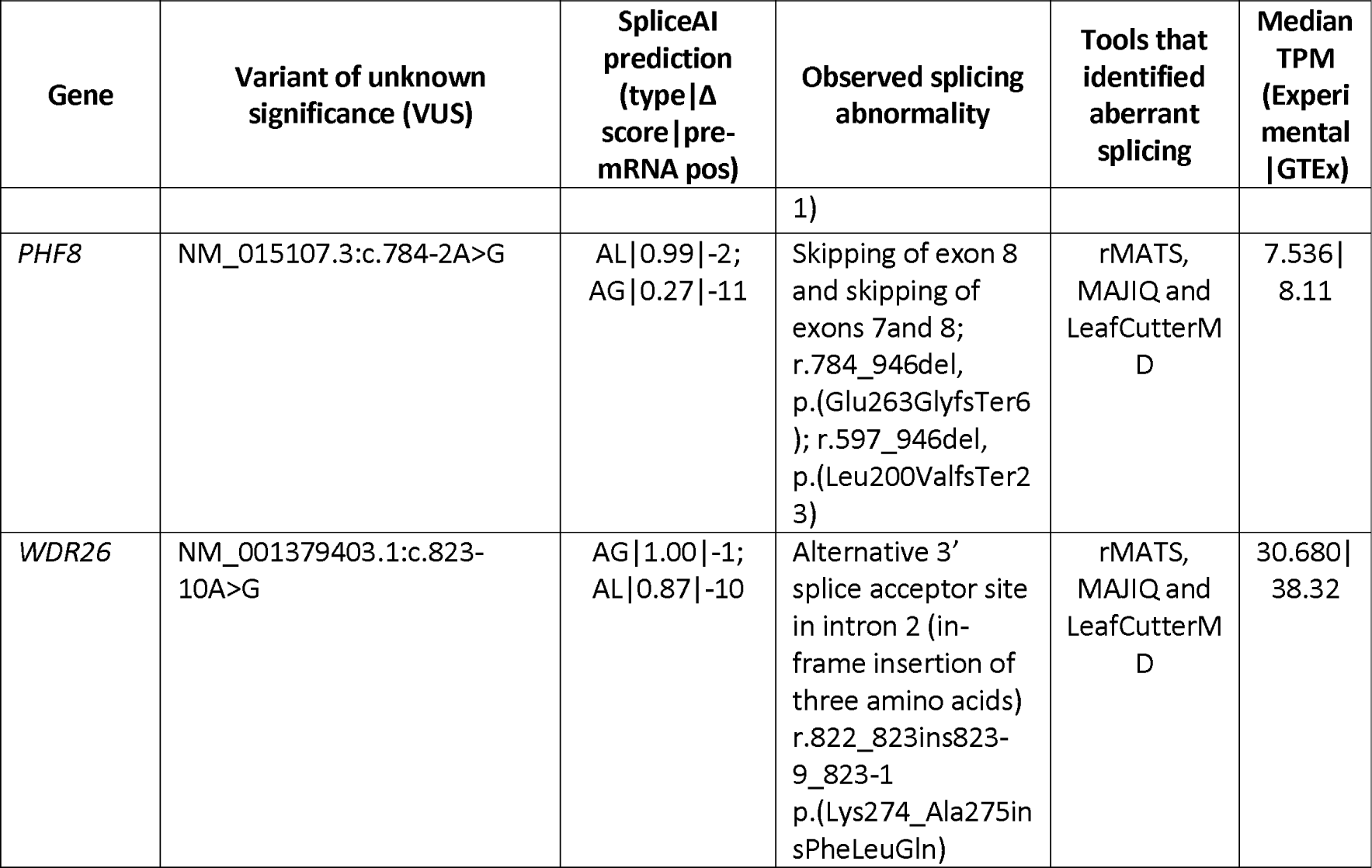
Variants of unknown significance (VUSs) for which aberrant splicing (AS) was observed in IGV. SpliceAI Lookup scores are indicated stating if donor loss (DL), donor gain (DG), acceptor loss (AL) or acceptor gain (AG) is predicted. Events that were identified by the splicing tools but had an adjusted p-value > 0.05 are denoted with an asterisk (*).

The DKC1 variant (NM_001363.5:c.915+10 G>A) was initially not found to affect splicing using RT-PCR in our previous publication(44). However, once the effect could be seen using RNA-seq, it was possible to design targeted assays to confirm the findings on RT-PCR. Thus, initial detection was not achieved by RT-PCR but post-RNA-seq confirmation was possible. We ran the RNA seq data through different splicing tools to identify the best tool/s to use when assessing cases with no candidate variants. The splicing tools rMATS-turbo, MAJIQ, FRASER2 and LeafCutterMD each identified 11, 4, 4, and 2 of the AS events respectively. rMATS-turbo had the best sensitivity identifying 79% of the AS events, where 7 were events identified solely by this tool. The aberrant splicing effects of three variants, SF3B4 c.417C>T, UBR4 c.8488+3A>G and SMARCE1 c.8-4A>G, were consistently missed by all tools. The SF3B4 variant is predicted to affect splicing (SpliceAI Δ score = 0.37), however, this gene has a high GC content and low mappability in large regions of its exons. Admittedly, only two reads mapped to the new junction and there were seven reads with the mutant allele that did not show aberrant splicing. Nonetheless, this event was validated via RT-PCR [results previously reported(10)] and has also been characterised using a β-globin hybrid minigene assay(45). The UBR4 variant is predicted to cause a donor loss leading to intron retention (SpliceAI Δ score = 0.26). In IGV 46 reads with the mutant allele and loss of the donor site were observed, but event was not detected by any of the tools (**Figure S4**). Lastly, the SMARCE1 variant is predicted to cause an acceptor loss (SpliceAI Δ score = 0.22). Like the SF3B4 variant, it also has few (n=9) reads mapping to the new junction. All three events were validated via RT-PCR and in the case of the UBR4 intron retention with additional qPCR. Using the evidence from RNAseq and RT-PCR results we were able to identify patterns in the data that would help us determine potential thresholds and limitations when assessing cases with no VUSs. This included: 1) SpliceAI predictions showed high concordance (sensitivity and specificity of 94% and 91% respectively) with the RNAseq and RT-PCR results; 2) In general, for VUSs which caused aberrant splicing, the variant was present in the data and reads with the mutant allele did not show normal splicing; 3) The splicing tools were able to detect aberrant events with as low as 5 reads supporting a new junction and; 4) Intron retention has a higher probability of being missed compared to other aberrant splicing events.

### Splicing analysis in patients without candidate VUSs

The patient cohort without a candidate VUS was comprised of 33 individuals plus an additional 23 cases where the original candidate VUS had not been found to cause aberrant splicing. To identify new candidate events in these cases, we took a systematic approach to narrow down the results obtained from rMATS-turbo to a manageable number so these could be inspected manually in IGV. rMATS-turbo was the preferred tool as it had the highest sensitivity identifying aberrant splicing events ascertained in the patients with a VUS. rMATS-turbo identified an average of 3,578 (2,370–115,522) significant events (FDR < 0.05) per sample with an inclusion level greater than 0.2 or less than −0.2.

Our first approach used filtered VCF files obtained by the RNA-seq variant calling pipeline to extract AS events within 25 base pairs of a variant. This first filtering step reduced the mean number of aberrant splicing events identified ∼300 fold to an average of 12 events per sample. Inspection of all events in IGV led to the identification of two new variants and associated aberrant splicing events.

### Case 1 – S075 (NARS1): identification of a splice-altering variant in a child with undiagnosed global developmental delay

rMATS-turbo identified two AS events within the NARS1 gene. The first event was an alternative donor site within exon 13 and the second was retention of intron 13 (**Figure 2**). These events were linked to a heterozygous missense variant within exon 13 (NARS1 c.1460C>T) predicted to affect splicing (SpliceAI Δ score = 0.93) by creating a new donor site. Deleterious variants in NARS1 are associated with Neurodevelopmental disorder with microcephaly, impaired language, and gait abnormalities, which would be consistent with the patient’s phenotype(46,47). NARS1 pathogenicity is generally associated with biallelic deleterious variants, however a recent study by Manole and colleagues has shown that de novo variants, including a recurrent nonsense variant at the end of the protein can have a gain-of-function effect that alters normal protein function by interfering with the ATP-binding domain, crucial for enzymatic function(48). In this case the intron retention is predicted to lead to an out-of-frame transcript, while the new donor site is predicted to lead to an in-frame deletion of 19 amino acids, both affecting the ATP-binding domain.

**Figure 2.**
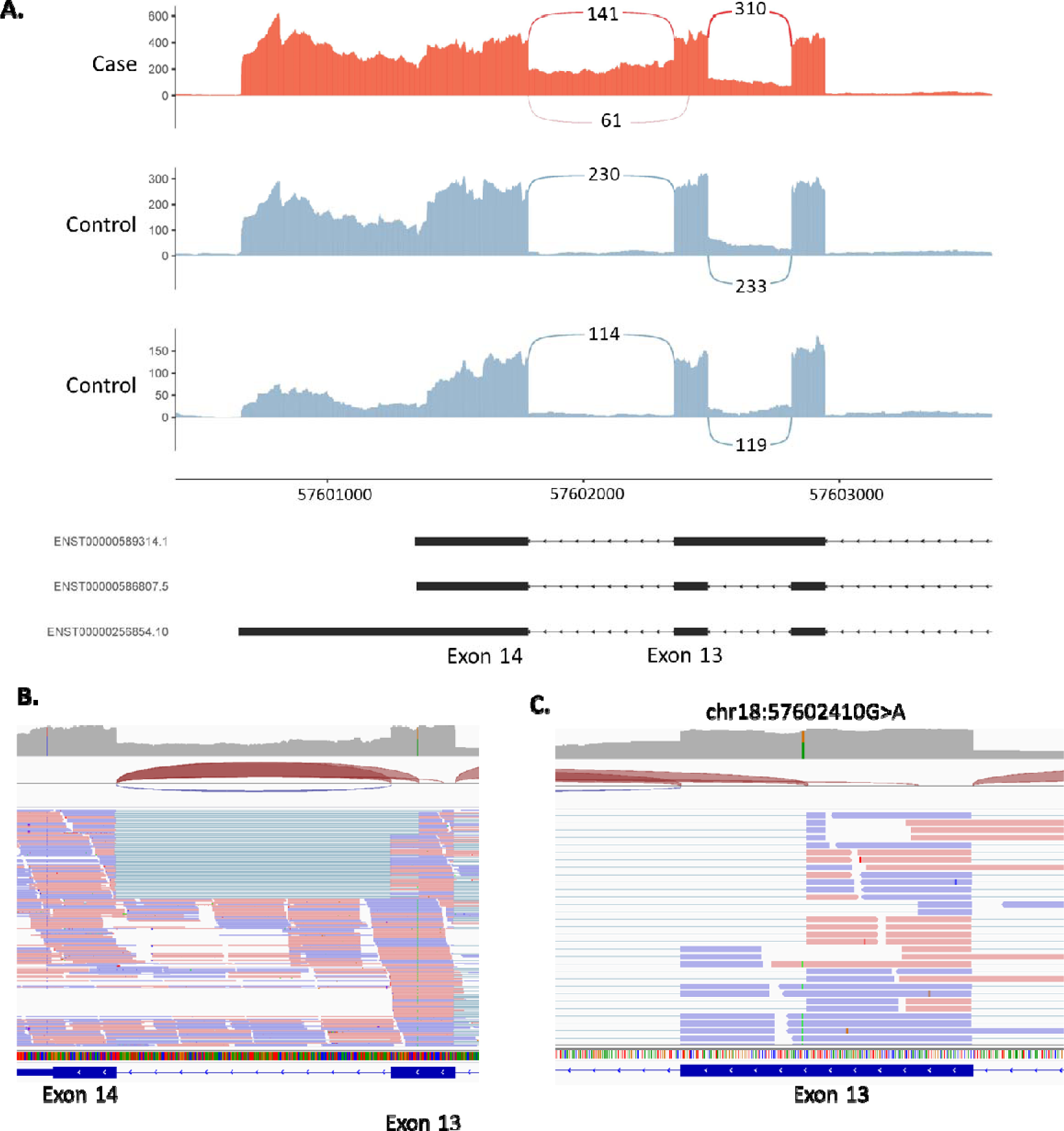
Alternative donor site and intron retention in *NARS1* gene. **A.** Sashimi plot of the proband and two controls of the alternative donor and intron retention region in *NARS1.* For the proband only (red track), we observed an alternative donor site in exon 13 as well as intron 13 retention. **B.** IGV screenshot of coverage across exons 13 and 14. **C.** Close up of *NARS1* c.1460C>T variant, a deep exonic variant predicted to affect splicing by creating a new donor site within exon 13. IGV images all derived from proband RNA-seq data.

### Case 2-S047 (ARFGEF1): inclusion of a cryptic exon in a child with undiagnosed developmental delay

rMATS-turbo identified an AS event within the ARFGEF1 gene associated with a deep intronic variant (chr8:67274263A>T, NM_006421.5:c.1337+1713T>A). This particular case was originally referred for analysis of a VUS (NM_138927.4:c.1160C>T), which after assessment in IGV was not observed to cause aberrant splicing. The ARFGEF1 variant is not predicted to affect splicing (SpliceAI Δ score = 0.0), however, the sequencing data shows the creation of a new acceptor and donor site within intron 9 suggesting the inclusion of a cryptic exon (**Figure 3**). Inclusion of this cryptic exon would result in an out-of-frame insertion of 186 nucleotides p.(Ser447PhefsTer19).

**Figure 3.**
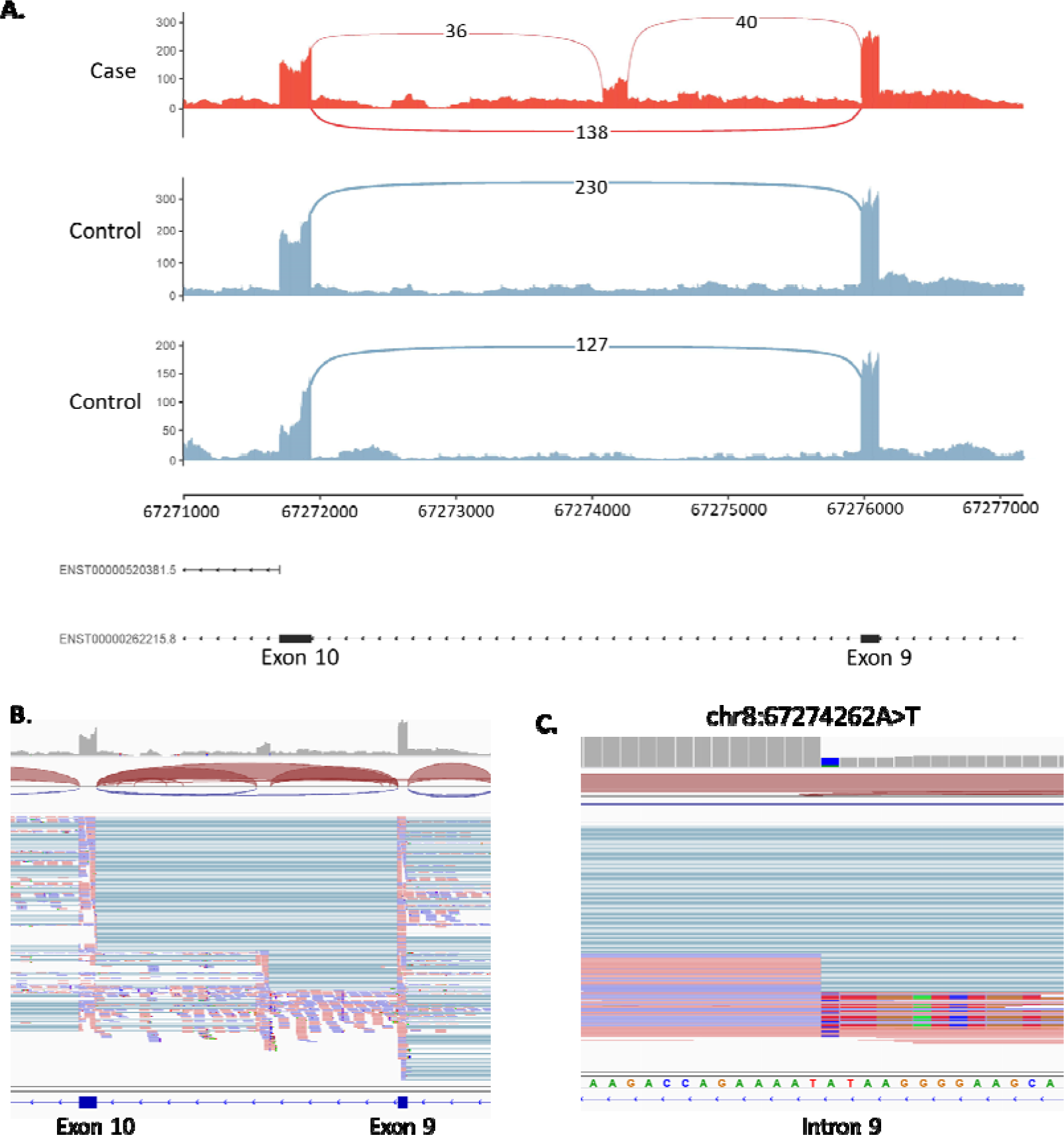
Activation of cryptic exon caused by intronic variant in *ARFGEF14* gene. **A.** Sashimi plot of the proband and two controls of the *ARFGEF14* region of interest. For the proband only (red track), two novel splice junctions can be seen suggesting the activation of a cryptic exon in intron 9. **B.** IGV screenshot of coverage across region of interest. **C.** Close up of chr8:67274262A>T variant. IGV images all derived from proband RNA-seq data.

Our second filtering strategy was a phenotype to genotype approach. Using the phenotype information available, results from the splicing tools were filtered using the appropriate Genomic Medicine Service (GMS) gene panels. This strategy led to the identification of one new candidate variant and associated aberrant splicing event.

### Case 3 – S076 (AP4E1): Identification of cryptic exon inclusion and a second frameshift variant in a child with undiagnosed hypotonia

The hypotonic infant GMS panel (v18.1) was applied to rMATS-turbo results, which identified activation of a pseudoexon within intron 1 of AP4E1 involving use of one alternative splice acceptor site and two alternative donor sites (**Figure 4**). The two resulting transcripts are predicted to be out of frame, leading to an insertion of 142 and 38 nucleotides. These events were associated with an intronic variant (chr15:50911536G>A, NM_007347.5:c.151-542G>A) weakly predicted to affect splicing. SpliceAI delta scores were 0.11 and 0.09 for acceptor gain (−32bp) and donor gain (5bp) respectively, but these were just below the 0.2 cut-off. However, analysis of the mutated sequence using ESEfinder predicts that the G>A base transition identified at this position may act as an exonic splicing enhancer through the creation of a binding site for splicing factors SC35 (SRSF2) and/or SRp40 (SRSF5)(49). This event was not picked up by the first method, as the variant was filtered out due to stringent quality thresholds [genotype quality (GQ) < 16; variant had a GQ of 6] required to manage noise when calling variants in RNA-seq data. Considering this gene has a biallelic mode of inheritance, we interrogated the rest of the gene for a second deleterious event and found a heterozygous single-nucleotide deletion in exon 6, NM_007347.5:c.567del, p.(Leu190TrpfsTer43), predicted to lead to an out-of-frame transcript (**Figure 4C**). Further testing confirmed that the variants are biparental. Biallelic variants in AP4E1 are associated with spastic paraplegia type 51, which is consistent with the phenotype information we have available for the proband.

**Figure 4.**
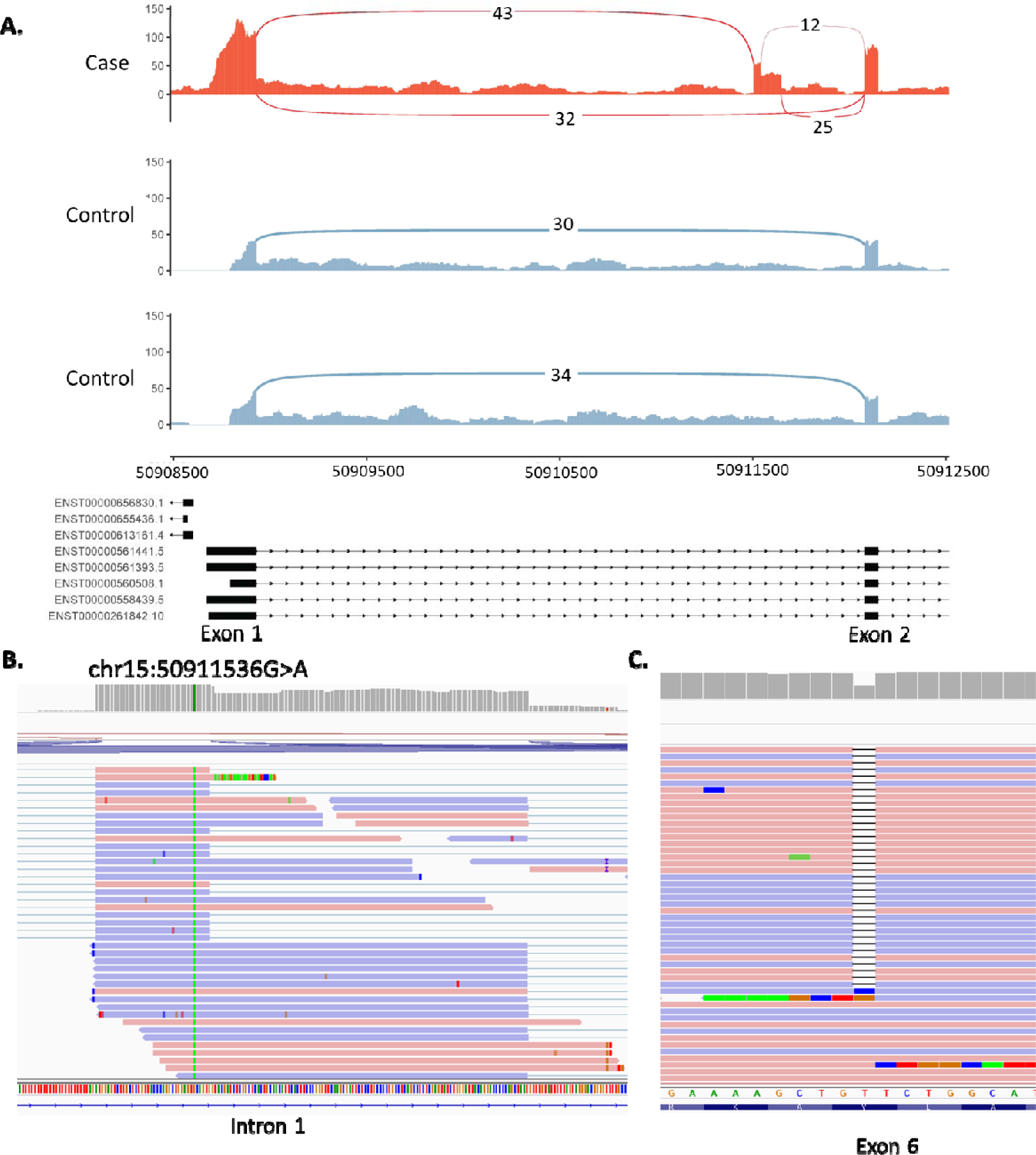
Activation of cryptic exon caused by intronic variant in the *AP4E1* gene. **A.** Sashimi plot of the proband and two controls of the *AP4E1* region of interest. For the proband only (red track), three novel splice junctions can be seen suggesting the activation of a cryptic exon in intron 1. **B.** Close up of NM_007347.5:c.151-542G>A variant in IGV. **C.** Heterozygous single nucleotide deletion observed in exon 6 (chr15:50929032delT). IGV images all derived from proband RNA-seq data.

All four variants identified (NM_006421.5:c.1337+1713T>A, NM_006421.5:c.1337+1713T>A, NM_007347.5:c.151-542G>A, and NM_007347.5:c.567del) were Sanger confirmed by the corresponding genetics laboratory subsequently generating three new diagnostic candidates across these 56 patients.

### Gene expression outlier analysis with OUTRIDER

Once aberrant splicing was systematically assessed we investigated if gene expression profiles would 1) generate new diagnostic candidates and 2) whether expression outliers correlated with aberrant splicing. OUTRIDER was run across the entire cohort excluding sample S017 for which only 16% of reads were uniquely mapping (n=86) and identified 175 gene expression outliers (FDR < 0.05) across 39 samples. Of the 39 samples that had expression outliers 16 were cases with a VUS, 18 were cases without a VUS and 5 were cases with known molecular diagnosis. Ten cases within our cohort had known chromosome microdeletions previously identified through microarray analysis (3 cases with known diagnosis and 7 with unknown diagnosis). In 5/10 of these cases, OUTRIDER identified genes with significantly lower expression which overlapped the deleted regions previously identified (**Figure 5**). For the 16 cases which had a VUS, none of the outliers identified matched the gene in which the VUS was found. A deeper analysis of the results did show that there were two variants, NM_001363.5(DKC1):c.915+10 G>A and NM_022356.4(P3H1):c.1224-80G>A, causing aberrant splicing for which the expression rank of the gene in which the VUS was found was 1 [lowest expression in the whole cohort] but was not significant after correction for multiple testing (**Figure S5**). The OUTRIDER gene p-value before correction was 0.0002 (z-score = −3.76) and 0.0009 (z-score= −3.31) for DCK1 and P3H1 respectively.

**Figure 5.**
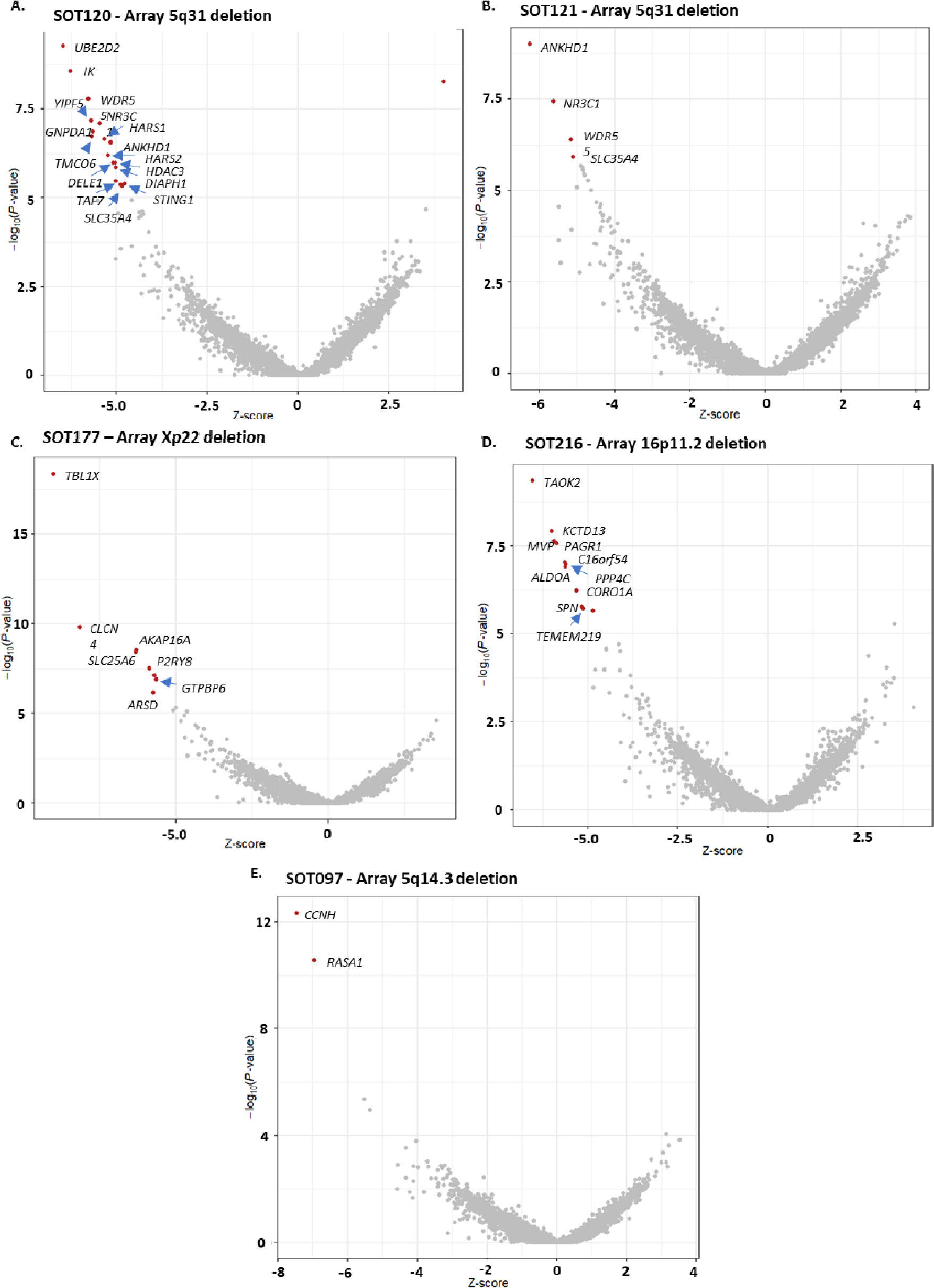
RNA aberrant expression detection with OUTRIDER. **A.** Volcano plot for sample SOT120, proband with an array 5q31 deletion. OUTRIDER identified 18 significant gene expression outliers (red) of which 17 fell within 5q31. **B.** Volcano plot for sample SOT121, proband with an array 5q31 deletion. OUTRIDER identified 4 significant gene expression outliers (red) of which 17 fell within 5q31. **C.** Volcano plot for sample SOT177, proband with Xp22 deletion. OUTRIDER identified 8 significant gene expression outliers (red) of which 7 fell within Xp22. **D.** Volcano plot for sample SOT216, proband with 16p11.2 deletion. OUTRIDER identified 11 significant gene expression outliers (red) of which 10 fell within deleted 16p11.2 region. **E.** Volcano plot for sample SOT097, proband with 5q14.3 deletion. OUTRIDER identified 2 significant gene expression outliers (red) both of which fell within the deleted region.

While gene expression profiles did not generate new diagnostic candidates, OUTRIDER results did lead to further investigation of one of the analysed cases (SOT177) to confirm skewed X-inactivation. This individual was a female child with developmental delay and dysmorphic features. Chromosome microarray analysis had identified a de novo 10.2 Mb deletion of Xp22.33p22.2. However, this copy number variant was classified as a VUS owing to the child being female and the assumption that the X chromosome carrying the deletion would be preferentially inactivated. Standard DNA-based X-inactivation testing proved uninformative in this case, but further primer sets showed unilateral inactivation. Trio whole-genome sequencing was subsequently undertaken to further seek a potential cause for the patient’s condition. No candidate variant was identified. However, it was possible to use parental SNP data to determine that the Xp deletion had occurred on the paternal X chromosome (**Figure S6**). Analysis of 9 additional heterozygous expressed SNPs in the patient’s RNA-seq data from loci across both arms of the X chromosome also revealed monoallelic paternal expression of X-linked genes (**Figure 6**). This therefore confirms complete skewing of X-inactivation towards the paternally inherited X-chromosome carrying the 10.2 Mb deletion. The cause of this extreme skewing currently remains unknown, as no candidates were found on the maternal X. However, the deletion is now thought to be causative for the patient’s presenting phenotype, resulting in a functional nullisomy for all genes in the deletion region that are subject to X inactivation.

**Figure 6.**
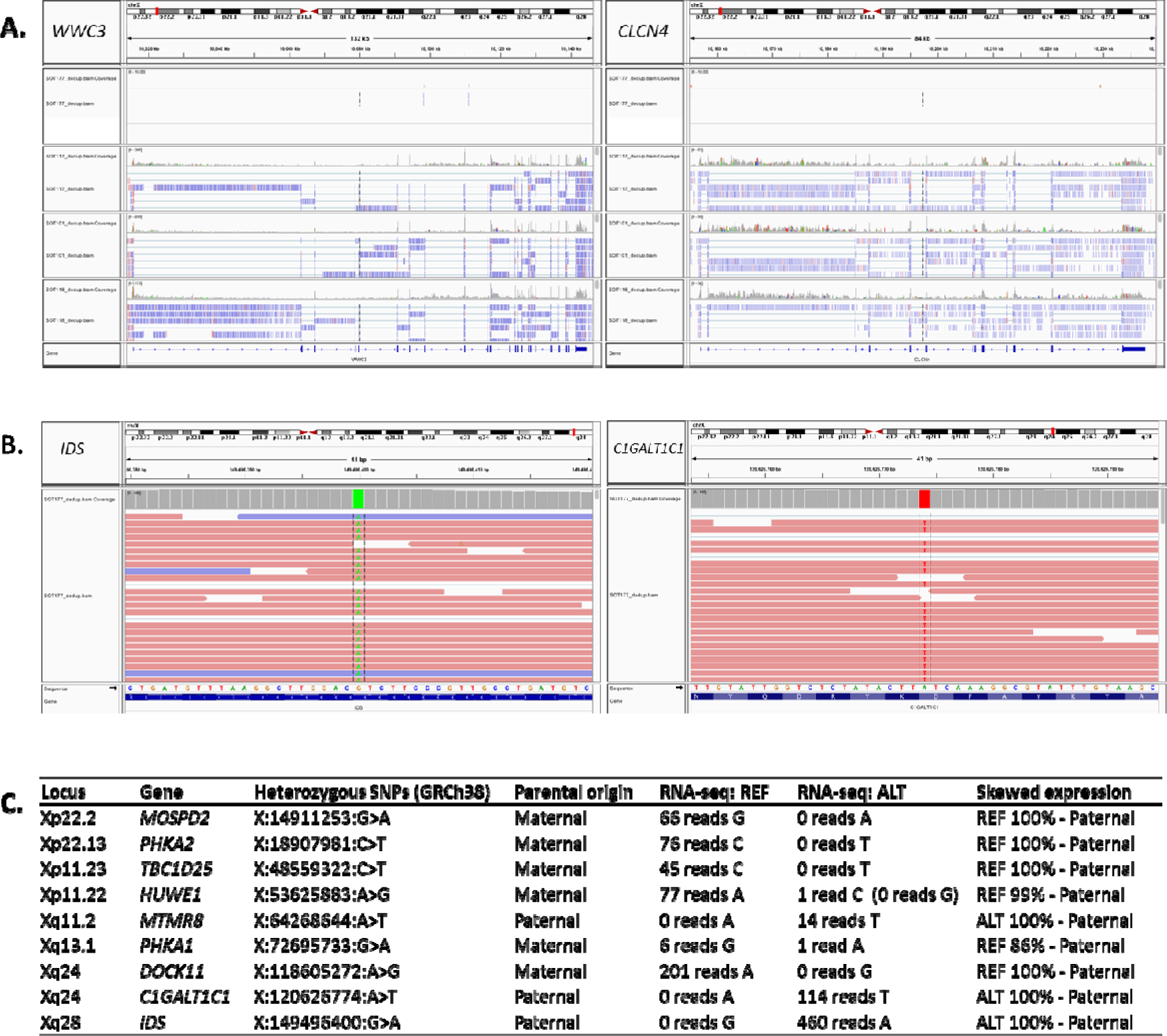
RNA-seq confirms skewed X-inactivation. **A.** IGV screenshots of RNA-seq data from *WWC3* and *CLCN4*, which lie within the Xp22.33p22.2 deletion. The patient’s sample (top track) shows no RNA-seq coverage compared to controls. **B.** IGV screenshots of RNA-seq data for heterozygous SNPs illustrating lack of maternal allele expression. **C**. Table of expression of selected heterozygous X-linked SNPs from across the X chromosome confirming skewing towards the paternal X.

## Discussion

In this work we have systematically assessed patients with no candidate VUSs in clinically relevant genes identifying three new diagnostic candidates and one additional diagnosis through analysis of expression profiles. We show that it is possible to make diagnoses using just RNA-seq in patients without a candidate VUS as well as classify VUSs using blood-based RNA-seq and RT-PCR to uplift diagnostic yield in rare disease patients. This work displays the variety of events that can be picked up using RNA-seq (i.e. deep intronic and exonic variants, complex splicing abnormalities, deletions, skewed x-inactivation) highlighting the wide range of applications this technology can have in the clinical setting.

### Splicing analysis in patients with VUSs

High-throughput blood-based RNA sequencing allowed us to evaluate the effect on splicing of 37/52 VUSs across 48 patients in clinically relevant genes. 38% of assessed VUSs (n=14) caused aberrant splicing detectable by RNA-seq, helping to clarify variant interpretation and provide supporting evidence of pathogenicity(50). For the 15/52 VUSs in which splicing could not be assessed using RNA-seq, the corresponding gene was not expressed in blood. In comparison to RNA-seq, RT-PCR proved to be more sensitive allowing us to assess 41 VUSs and confirmed a further four likely pathogenic AS events, meaning that in total 35% of VUSs in this cohort (44% of those that could be adequately assayed) were found to affect splicing (**see Table S1**). These figures are in concordance with SpliceAI predictions which had a sensitivity and specificity of 94% and 91% respectively. The increase in sensitivity of RT-PCR can be attributed to the targeted approach allowing amplification of AS events in lowly expressed genes(51), as well as amplification of AS events with low inclusion levels (in some cases accounting for NMD). For the nine cases where neither RNA-seq nor RT-PCR was able to resolve the VUS, such variants would need to be assessed with other tissue types or via alternative methods such as minigene analysis or potentially using animal models should the collection of an appropriate or adequately representative tissue not be feasible(51). The MRSD tool was used to predict the minimum required sequencing depth for genes of interest and was found to be very conservative. From our empirical data, genes with GTEx whole-blood TPM values of 5 or above are likely to be assessable for splicing analysis using the RNA-seq parameters employed in this study, while genes with TPM values down to 0.9 may be assessable by RT-PCR. A TPM threshold of 5 would correspond to 1104/3113 (35%) of genes listed in the UK Genomic Medicine Service’s PanelApp(31) list of disease genes, while a threshold of 0.9 would include 1866/3113 (60%), see **Figure S7**. Based on our analysis, we recommend that RT-PCR should be the first-choice test to assess VUSs in genes with low expression in blood such as BRCA1, BRCA2 and FBN1. In some instances, informative RT-PCR results can be obtained even in genes reported to have a TPM value of zero in GTEx(10). However, in most other cases RNA-seq is likely to prove more advantageous as a first line test. RNA-seq can identify splicing events with more granularity, particularly when new AS events entail only one or a few nucleotides. Furthermore, with RNA-seq we can quantify splice isoforms, identify expression outliers, and most importantly we can look at aberrant splicing events without prior expectation of what the causal variants may be. Case 4 (SOT247) highlights just how useful having transcriptome wide data is, as this patient was referred with a VUS that did not cause aberrant splicing, but the ability to look at the entire transcriptome led to the identification of a likely disruptive splicing event in a different gene.

While the VUSs in this cohort were enriched for variants affecting splicing, these were clinically identified VUSs for which clarification of pathogenicity was sought by clinicians, highlighting the need for this type of tests to be integrated into clinical practice. Overall, we were able to assess 83% of VUSs (RNA-seq and RT-PCR combined), confirming the utility of blood as a suitable tissue for validating aberrant splicing in rare disease patients.

### Identification of splicing events linked to VUSs by different splicing tools

While we did not set out to benchmark a comprehensive selection of splice junction detection tools, we did however want to establish if widely used tools could be used to detect aberrant splicing in rare disease patients as datasets used in previous benchmarking studies were not comparable to ours(52–54). Using the 14 cases which had aberrant splicing linked to known VUSs, rMATS-turbo had the highest sensitivity followed by FRASER2, MAJIQ and then LeafcutterMD. FRASER2 and LeafCutterMD were the two tools developed for outlier splicing detection and therefore it was unexpected LeafCutterMD had the lowest sensitivity. There were three variants which were consistently missed by the splicing tools (SF3B4 c.417C>T, UBR4 c.8488+3A>G and SMARCE1 c.8-4A>G) and is likely that the low number of reads covering the variants within SF3B4 and SMARCE1 is the reason the splicing tools are not picking up these AS events. We suspect that the low number of reads supporting the new splicing events are due to nonsense mediated decay or leaky splicing, however in these two cases, there weren’t any coding SNPs to confirm NMD. Long-read may help with this partially but for short-read data, determining the difference between partial/leaky splicing and NMD, or possible feedback effects on decreased transcription, will require new analytical methods. Furthermore, with regards to the intron retention in UBR4, it is unclear why the tools are not identifying this event as there is strong supporting evidence (**Figure S4**).

### Splicing analysis in patients without VUSs

In patients with no prior VUS, the sheer number of significant events resulting from splicing tools create a challenge in terms of identifying new potential aberrant splicing events that could be linked to the patients’ conditions. However, we were able to filter these down to a manageable number and identify new likely disruptive aberrant splicing events albeit with strict filtering criteria, rendering it likely genuine events were missed.

Out of a total of 56 cases without a previously identified VUS or with a VUS but with no aberrant splicing observed, our RNA-seq analysis identified three cases with relevant splicing alterations and one case with skewed X-inactivation, suggesting a potential diagnostic uplift rate of 7%. This is an important untapped group of variants with few established high-throughput methods of analysis in these types of cohorts(55). We thereby demonstrate that it is possible to identify new candidate diagnoses and splicing events in patients with no prior candidate sequence variants, although with a much lower yield than if a VUS were previously identified. Two of the four new diagnostics candidates were caused by deep intronic variants, regions of the genome often overlooked in genomic investigations and where it is difficult to predict functional effects. If prediction algorithms are to be used for prioritisation of variants, these may need to be tailored by genomic region, such as having a more permissive SpliceAI score threshold for deep intronic variants. As demonstrated by the activation of a cryptic exon caused by a deep intronic variant in the ARFGEF1 gene whose SpliceAI delta score was just below the widely used 0.2 cut-off.

The low diagnostic yield in patients without a candidate VUS could be attributed to a number of factors: 1) The patient could have a variant affecting splicing in a gene that is not expressed in blood or there is a tissue specific impact that is not present in blood; 2) The molecular cause of the disease does not affect splicing; 3) The tools are not able to identify these events with high confidence (e.g. aberrant isoform is undergoing nonsense mediated decay or difficult to align); 4) Performance of the tools is variable, some events are picked up better than others (e.g. exon skipping compared to intron retention); and 5) The variant could have been filtered out.

We were limited to using RNA for variant calling as there was no matching DNA sequencing available for most cases in this study. Consequently, the high number of false positive variant calls led to strict filtering criteria where only SNVs were inspected and thus events caused by indels will have been missed. RNA sequencing may not be ideal for variant calling as it generates high numbers of false positive calls compared to DNA sequencing due to both biological and technical differences. Nonetheless, the use of gene panels to restrict results from the splicing tool (rMATS-turbo) did recover a variant that had been excluded due to harsh filters. This approach does limit the analysis by restricting to known disease genes and relies on having robust phenotypic information. If matched whole genome sequencing data and detailed phenotypic information were available, integration of this data would likely increase events identified in this patient subgroup and potentially increase diagnostic yield.

### Gene expression analysis

OUTRIDER was able to detect half of the known microdeletions in the cohort, but it did not detect significant alterations in gene expression in genes which did show aberrant splicing. There were only two instances where the VUS gene was ranked first (lowest expression for the whole cohort) and although neither passed the significance threshold, this suggest that there is a likely decrease of normal transcripts and perhaps decreasing the number of genes tested (i.e. OMIM or PanelApp genes) could increase the number of significant events identified as well as decreasing the amount of false positives. This finding also indicates that abnormal splicing is not necessarily associated with a significant reduction in gene expression, at least in blood, and over-reliance on such expression changes for the identification of splicing abnormalities is unlikely to have reliable sensitivity. This is particularly interesting as we would expect many of the splicing abnormalities to shift the reading frame and therefore undergo nonsense mediated decay (NMD) significantly decreasing the abundance of the transcript. This lack of change in expression of genes with aberrant splicing could be a reflection of biological mechanisms indicating that the impact of NMD is not as effective at depleting aberrant transcripts or it could be due to technical factors such as limited sensitivity of the tools as mentioned previously; the impact of NMD is not very pronounced in blood-based RNA-seq and tissue specific RNA-seq is required; and/or the targeting methodologies bias the type of transcripts and number of transcripts we observe. Furthermore, whole blood has been shown to have high variability in gene expression profiles particularly when compared to skin fibroblasts(16). Some studies suggest that fibroblast RNA enables the investigation of a more comprehensive set of genes than whole blood and that this is likely the better tissue for detecting clinically relevant differences in gene expression(12,14,32). While blood-based RNA analysis may not be optimal, it does offer a number of benefits over fibroblasts. It is more routinely sampled, less invasive to obtain and does not require cell culture prior to testing, meaning it is faster to obtain and analyse and has lower requirements in terms of specialised knowledge and facilities.

### Therapeutic applications

Accurate diagnoses facilitate appropriate clinical management, accurate genetic counselling and informed reproductive decision making, but in some cases, there would be the potential for bespoke RNA-targeted therapies to be designed to correct a given splicing abnormality and slow or halt the progression of an individual’s disease. Cases such as that of the AP4E1 cryptic exon inclusion variant highlighted in this study may be especially suitable targets in this regard, on account of the gradual neurodegenerative nature of the associated condition and the known efficacy of other antisense oligonucleotide therapies delivered to the central nervous system such as nusinersen(56,57). Notwithstanding the substantial challenges and barriers facing development of such bespoke therapeutics, precedent does exist for n=1 oligonucleotide therapies(58). The utility of RNA-seq in being able to identify these types of variants means that an effective personalised medicine healthcare system will benefit from having access to RNA transcriptomics within the diagnostic clinical setting.

## Conclusion

To our knowledge, this study is the first to incorporate variant calling data from RNA-seq to results from splicing tools to identify new diagnostic candidates in rare disease. While the diagnostic uplift is modest in patients with no known candidate variants in clinically relevant genes, our analyses suggest at least one third of patients with rare disorders could benefit from the increased diagnostic yield offered by RNA-seq by providing additional functional evidence for VUSs. When considering analysis of RNA, RT-PCR should be the first-choice test to assess VUSs in genes with low expression, but high throughput RNA sequencing is more advantageous as a first line test. Overall, we were able to validate splicing abnormalities in 35% [18/51] of patients with a VUS and identified four new diagnoses by detecting novel AS and expression events in patients with no candidate sequence variants, giving an overall uplift in diagnostic yield of 7% [4/56] in this subset of patients. We believe, that RNA-seq should be considered as a complementary tool in genetic testing to uplift diagnostic yield in cohorts of patients with rare disorders particularly when integrated with other omic data.

## List of abbreviations

VUS: Variant of unknown significance

IGV: Integrative Genomics Viewer

RNA-seq: RNA sequencing

AS: Aberrant splicing

RT-PCR: Reverse transcription PCR

## Declarations

### Ethics approval and consent to participate

Participants were enrolled into the University of Southampton’s Splicing and Disease study with appropriate ethical approval (REC 11/SC/0269, IRAS 49685, ERGO 23056).

## Consent for publication

Not applicable

## Availability of data and materials

The datasets generated and/or analysed during the current study are not publicly available as ethics approval and consent agreements allow us to share non-identifiable patient data and analysis data only, as such, we cannot provide BAM or VCF files. The code generated during this study can be found in GitHub [https://github.com/carojoquendo/RNA_splicing_and_disease].

## Author contributions

Conceptualization, D.B, A.G.L.D, J.L; Data Curation C.J.O, J.L; Formal Analysis C.J.O, A.G.L.D, J.L, H.A.W; Funding Acquisition D.B; Investigation C.J.O, A.G.L.D, J.L, H.A.W, W.R, D.J.B, D.H, N.S.T; Methodology C.J.O, A.G.L.D, J.L, H.A.W; Project administration, D.B; Resources, D.B, A.G.L.D; Software, C.J.O, J.L, W.R; Supervision D.B; Validation, H.A.W, A.G.L.D, D.H;D.J.B; Visualization C.J.O; Writing – Original Draft, C.J.O, A.G.L.D, Writing – Review & Editing; C.J.O, A.G.L.D, J.L, D.J.B, D.H, N.S.T, D.B.

## Data Availability

All data produced in the present study are available upon reasonable request to the authors

https://github.com/carojoquendo/RNA_splicing_and_disease

## Acknowledgements

The authors thank all patients and families taking part in this research and all clinicians involved. The authors acknowledge the use of the IRIDIS High Performance Computing Facility, and associated support services at the University of Southampton, in the completion of this work.

## Additional files

**Figure S1:** PDF: Overview of RNA sequencing data. Additional figure detailing, distribution of uniquely mapping reads, total number of junctions, correlation of experimental TPM values versus GTEx values as well as clustering of samples based on transcriptomic profiles.

**Figure S2:** PDF: Median transcript per million (TPM) values in genes with a variant of uncertain significance (VUS) across the four batches. Bar plot comparing TPM values across each batch in genes of interest.

**Figure S3:** PDF: Effects of globin depletion on gene expression. Figure showing correlation and comparison of TPM values between globin depleted and non-depleted batches. **Figure S4**: PDF: Intron retention caused by intronic variant in UBR4. Sashimi plot and RNA-seq IGV screenshot of aberrant splicing observed in patient with UBR4 variant.

**Figure S5:** PDF: Gene rank plots for DKC1 and P3H1. Dot plot showing OUTRIDER expression rank for DKC1 and P3H1.

**Figure S6:** PDF: Trio whole genome sequencing confirms paternal chromosomal origin of a Xp22.33p22.2 deletion. SNP data within the deleted region of female patient with Xp22.33p22.2 deletion of 10.2 Mb.

**Figure S7:** PDF: GTEx blood TPM values across selected different gene panels available via the UK Genomic Medicine Service. Breakdown of GTEx blood TPM values for selected panels available in the UK Genomic Medicine Service.

**Table S1:** xlsx: Detailed list of samples and variants assessed using RNA-seq and RT-PCR.

## References

1. Ewans LJ, Minoche AE, Schofield D, Shrestha R, Puttick C, Zhu Y, et al. Whole exome and genome sequencing in mendelian disorders: a diagnostic and health economic analysis. Eur J Hum Genet. 2022 Oct 1;30(10):1121–31.

2. Nurchis MC, Altamura G, Riccardi MT, Radio FC, Chillemi G, Bertini ES, et al. Whole genome sequencing diagnostic yield for paediatric patients with suspected genetic disorders: systematic review, meta-analysis, and GRADE assessment. Arch Public Health. 2023 May 25;81(1):93.

3. 100,000 Genomes Pilot on Rare-Disease Diagnosis in Health Care — Preliminary Report. N Engl J Med. 2021 Nov 11;385(20):1868–80.

4. Álvarez-Mora MI, Sánchez A, Rodríguez-Revenga L, Corominas J, Rabionet R, Puig S, et al. Diagnostic yield of next-generation sequencing in 87 families with neurodevelopmental disorders. Orphanet J Rare Dis. 2022 Feb 19;17(1):60.

5. Cummings BB, Marshall JL, Tukiainen T, Lek M, Donkervoort S, Foley AR, et al. Improving genetic diagnosis in Mendelian disease with transcriptome sequencing. Sci Transl Med. 2017 Apr 19;9(386).

6. Kremer LS, Bader DM, Mertes C, Kopajtich R, Pichler G, Iuso A, et al. Genetic diagnosis of Mendelian disorders via RNA sequencing. Nat Commun. 2017 Jun 12;8:15824.

7. Frésard L, Smail C, Ferraro NM, Teran NA, Li X, Smith KS, et al. Identification of rare-disease genes using blood transcriptome sequencing and large control cohorts. Nat Med. 2019 Jun;25(6):911–9.

8. Gonorazky HD, Naumenko S, Ramani AK, Nelakuditi V, Mashouri P, Wang P, et al. Expanding the Boundaries of RNA Sequencing as a Diagnostic Tool for Rare Mendelian Disease. Am J Hum Genet. 2019 May 2;104(5):1007.

9. Hamanaka K, Miyatake S, Koshimizu E, Tsurusaki Y, Mitsuhashi S, Iwama K, et al. RNA sequencing solved the most common but unrecognized NEB pathogenic variant in Japanese nemaline myopathy. Genet Med Off J Am Coll Med Genet. 2019 Jul;21(7):1629–38.

10. Wai HA, Lord J, Lyon M, Gunning A, Kelly H, Cibin P, et al. Blood RNA analysis can increase clinical diagnostic rate and resolve variants of uncertain significance. Genet Med Off J Am Coll Med Genet. 2020 Jun;22(6):1005–14.

11. Colin E, Duffourd Y, Chevarin M, Tisserant E, Verdez S, Paccaud J, et al. Stepwise use of genomics and transcriptomics technologies increases diagnostic yield in Mendelian disorders. Front Cell Dev Biol [Internet]. 2023;11. Available from: https://www.frontiersin.org/articles/10.3389/fcell.2023.1021920

12. Yépez VA, Gusic M, Kopajtich R, Mertes C, Smith NH, Alston CL, et al. Clinical implementation of RNA sequencing for Mendelian disease diagnostics. Genome Med [Internet]. 2022;14(1). Available from: https://www.scopus.com/inward/record.uri?eid=2-s2.0-85127470335&doi=10.1186%2fs13073-022-01019-9&partnerID=40&md5=2f051e53e887cc2ce85156235eb677e1

13. Dekker J, Schot R, Bongaerts M, Valk WG de, Veghel-Plandsoen MM van, Monfils K, et al. Web-accessible application for identifying pathogenic transcripts with RNA-seq: Increased sensitivity in diagnosis of neurodevelopmental disorders. Am J Hum Genet. 2023;110(2):251–72.

14. Lee H, Huang AY, Wang LK, Yoon AJ, Renteria G, Eskin A, et al. Diagnostic utility of transcriptome sequencing for rare Mendelian diseases. Genet Med Off J Am Coll Med Genet. 2020 Mar;22(3):490–9.

15. Maddirevula S, Kuwahara H, Ewida N, Shamseldin HE, Patel N, Alzahrani F, et al. Analysis of transcript-deleterious variants in Mendelian disorders: implications for RNA-based diagnostics. Genome Biol. 2020 Jun 17;21(1):145.

16. Murdock DR, Dai H, Burrage LC, Rosenfeld JA, Ketkar S, Müller MF, et al. Transcriptome-directed analysis for Mendelian disease diagnosis overcomes limitations of conventional genomic testing [Internet]. Vol. 131, Journal of Clinical Investigation. 2021. Available from: https://www.scopus.com/inward/record.uri?eid=2-s2.0-85098878187&doi=10.1172%2fJCI141500&partnerID=40&md5=5b4afe46dd2a2aa7cd520d169897d88d

17. Frankish A, Carbonell-Sala S, Diekhans M, Jungreis I, Loveland JE, Mudge JM, et al. GENCODE: reference annotation for the human and mouse genomes in 2023. Nucleic Acids Res. 2023 Jan 6;51(D1):D942–9.

18. Dobin A, Davis CA, Schlesinger F, Drenkow J, Zaleski C, Jha S, et al. STAR: ultrafast universal RNA-seq aligner. Bioinforma Oxf Engl. 2013 Jan 1;29(1):15–21.

19. Robinson JT, Thorvaldsdóttir H, Winckler W, Guttman M, Lander ES, Getz G, et al. Integrative genomics viewer. Nat Biotechnol. 2011 Jan;29(1):24–6.

20. Garrido-Martín D, Palumbo E, Guigó R, Breschi A. ggsashimi: Sashimi plot revised for browser-and annotation-independent splicing visualization. PLOS Comput Biol. 2018 Aug;14(8):1–6.

21. Mertes C, Scheller IF, Yépez VA, Çelik MH, Liang Y, Kremer LS, et al. Detection of aberrant splicing events in RNA-seq data using FRASER. Nat Commun. 2021 Jan 22;12(1):529.

22. Scheller IF, Lutz K, Mertes C, Yépez VA, Gagneur J. Improved detection of aberrant splicing using the Intron Jaccard Index [Internet]. medRxiv; 2023 [cited 2023 Nov 3]. p. 2023.03.31.23287997. Available from: https://www.medrxiv.org/content/10.1101/2023.03.31.23287997v1

23. Shen S, Park JW, Lu Z xiang, Lin L, Henry MD, Wu YN, et al. rMATS: robust and flexible detection of differential alternative splicing from replicate RNA-Seq data. Proc Natl Acad Sci U S A. 2014 Dec 23;111(51):E5593–5601.

24. Vaquero-Garcia J, Barrera A, Gazzara MR, González-Vallinas J, Lahens NF, Hogenesch JB, et al. A new view of transcriptome complexity and regulation through the lens of local splicing variations. eLife. 2016 Feb 1;5:e11752.

25. Jenkinson G, Li YI, Basu S, Cousin MA, Oliver GR, Klee EW. LeafCutterMD: an algorithm for outlier splicing detection in rare diseases. Bioinforma Oxf Engl. 2020 Nov 1;36(17):4609–15.

26. Van der Auwera GA, O’Connor BD. Genomics in the Cloud: Using Docker, GATK, and WDL in Terra [Internet]. O’Reilly Media; 2020. Available from: https://books.google.co.uk/books?id=vsXaDwAAQBAJ

27. Danecek P, Bonfield JK, Liddle J, Marshall J, Ohan V, Pollard MO, et al. Twelve years of SAMtools and BCFtools. GigaScience. 2021 Feb 16;10(2).

28. Jehl F, Degalez F, Bernard M, Lecerf F, Lagoutte L, Désert C, et al. RNA-Seq Data for Reliable SNP Detection and Genotype Calling: Interest for Coding Variant Characterization and Cis-Regulation Analysis by Allele-Specific Expression in Livestock Species. Front Genet. 2021;12:655707.

29. McLaren W, Gil L, Hunt SE, Riat HS, Ritchie GRS, Thormann A, et al. The Ensembl Variant Effect Predictor. Genome Biol. 2016 Jun 6;17(1):122.

30. Quinlan AR, Hall IM. BEDTools: a flexible suite of utilities for comparing genomic features. Bioinforma Oxf Engl. 2010 Mar 15;26(6):841–2.

31. Martin AR, Williams E, Foulger RE, Leigh S, Daugherty LC, Niblock O, et al. PanelApp crowdsources expert knowledge to establish consensus diagnostic gene panels. Nat Genet. 2019 Nov;51(11):1560–5.

32. Rowlands CF, Taylor A, Rice G, Whiffin N, Hall HN, Newman WG, et al. MRSD: A quantitative approach for assessing suitability of RNA-seq in the investigation of mis-splicing in Mendelian disease. Am J Hum Genet. 2022;109(2):210–22.

33. Putri GH, Anders S, Pyl PT, Pimanda JE, Zanini F. Analysing high-throughput sequencing data in Python with HTSeq 2.0. Bioinformatics. 2022 May 13;38(10):2943–5.

34. Piovesan A, Caracausi M, Ricci M, Strippoli P, Vitale L, Pelleri MC. Identification of minimal eukaryotic introns through GeneBase, a user-friendly tool for parsing the NCBI Gene databank. DNA Res Int J Rapid Publ Rep Genes Genomes. 2015 Dec;22(6):495–503.

35. Neph S, Kuehn MS, Reynolds AP, Haugen E, Thurman RE, Johnson AK, et al. BEDOPS: high-performance genomic feature operations. Bioinforma Oxf Engl. 2012 Jul 15;28(14):1919–20.

36. Patro R, Duggal G, Love MI, Irizarry RA, Kingsford C. Salmon provides fast and bias-aware quantification of transcript expression. Nat Methods. 2017 Apr;14(4):417–9.

37. R Core Team. R: A language and environment for statistical computing [Internet]. Vienna, Austria: R Foundation for Statistical Computing; 2021. Available from: https://www.R-project.org/

38. Soneson C, Love MI, Robinson MD. Differential analyses for RNA-seq: transcript-level estimates improve gene-level inferences. F1000Research. 2015;4:1521.

39. Brechtmann F, Mertes C, Matusevičiūtė A, Yépez VA, Avsec Ž, Herzog M, et al. OUTRIDER: A Statistical Method for Detecting Aberrantly Expressed Genes in RNA Sequencing Data. Am J Hum Genet. 2018 Dec 6;103(6):907–17.

40. The Genotype-Tissue Expression (GTEx) project. Nat Genet. 2013 Jun;45(6):580–5.

41. Amberger JS, Bocchini CA, Schiettecatte F, Scott AF, Hamosh A. OMIM.org: Online Mendelian Inheritance in Man (OMIM®), an online catalog of human genes and genetic disorders. Nucleic Acids Res. 2015 Jan;43(Database issue):D789-798.

42. Harrington CA, Fei SS, Minnier J, Carbone L, Searles R, Davis BA, et al. RNA-Seq of human whole blood: Evaluation of globin RNA depletion on Ribo-Zero library method. Sci Rep. 2020 Apr 14;10(1):6271.

43. Carithers LJ, Ardlie K, Barcus M, Branton PA, Britton A, Buia SA, et al. A Novel Approach to High-Quality Postmortem Tissue Procurement: The GTEx Project. Biopreservation Biobanking. 2015 Oct;13(5):311–9.

44. Wai HA, Lord J, Lyon M, Gunning A, Kelly H, Cibin P, et al. Blood RNA analysis can increase clinical diagnostic rate and resolve variants of uncertain significance. Genet Med. 2020 Jun 1;22(6):1005–14.

45. Cassina M, Cerqua C, Rossi S, Salviati L, Martini A, Clementi M, et al. A synonymous splicing mutation in the SF3B4 gene segregates in a family with highly variable Nager syndrome. Eur J Hum Genet EJHG. 2017 Feb;25(3):371–5.

46. Wang L, Li Z, Sievert D, Smith DEC, Mendes MI, Chen DY, et al. Loss of NARS1 impairs progenitor proliferation in cortical brain organoids and leads to microcephaly. Nat Commun. 2020 Aug 12;11(1):4038.

47. Manole A, Efthymiou S, O’Connor E, Mendes MI, Jennings M, Maroofian R, et al. De Novo and Bi-allelic Pathogenic Variants in NARS1 Cause Neurodevelopmental Delay Due to Toxic Gain-of-Function and Partial Loss-of-Function Effects. Am J Hum Genet. 2020 Aug 6;107(2):311–24.

48. Manole A, Efthymiou S, O’Connor E, Mendes MI, Jennings M, Maroofian R, et al. De Novo and Bi-allelic Pathogenic Variants in NARS1 Cause Neurodevelopmental Delay Due to Toxic Gain-of-Function and Partial Loss-of-Function Effects. Am J Hum Genet. 2020 Aug 6;107(2):311–24.

49. Cartegni L, Wang J, Zhu Z, Zhang MQ, Krainer AR. ESEfinder: A web resource to identify exonic splicing enhancers. Nucleic Acids Res. 2003 Jul 1;31(13):3568–71.

50. Richards S, Aziz N, Bale S, Bick D, Das S, Gastier-Foster J, et al. Standards and guidelines for the interpretation of sequence variants: a joint consensus recommendation of the American College of Medical Genetics and Genomics and the Association for Molecular Pathology. Genet Med. 2015 May 1;17(5):405–23.

51. Wai HA, Constable M, Drewes C, Davies IC, Svobodova E, Dempsey E, et al. Short amplicon reverse transcription-polymerase chain reaction detects aberrant splicing in genes with low expression in blood missed by ribonucleic acid sequencing analysis for clinical diagnosis. Hum Mutat. 2022 Jul;43(7):963–70.

52. Ding L, Rath E, Bai Y. Comparison of Alternative Splicing Junction Detection Tools Using RNA-Seq Data. Curr Genomics. 2017 Jun;18(3):268–77.

53. Mehmood A, Laiho A, Venäläinen MS, McGlinchey AJ, Wang N, Elo LL. Systematic evaluation of differential splicing tools for RNA-seq studies. Brief Bioinform. 2020 Dec 1;21(6):2052–65.

54. Jiang M, Zhang S, Yin H, Zhuo Z, Meng G. A comprehensive benchmarking of differential splicing tools for RNA-seq analysis at the event level. Brief Bioinform. 2023 Apr 5;bbad121.

55. Marwaha S, Knowles JW, Ashley EA. A guide for the diagnosis of rare and undiagnosed disease: beyond the exome. Genome Med. 2022 Feb 28;14(1):23.

56. Ebrahimi-Fakhari D, Teinert J, Behne R, Wimmer M, D’Amore A, Eberhardt K, et al. Defining the clinical, molecular and imaging spectrum of adaptor protein complex 4-associated hereditary spastic paraplegia. Brain. 2020 Oct 1;143(10):2929–44.

57. Pane M, Coratti G, Sansone VA, Messina S, Catteruccia M, Bruno C, et al. Type I spinal muscular atrophy patients treated with nusinersen: 4-year follow-up of motor, respiratory and bulbar function. Eur J Neurol. 2023 Mar 7;

58. Kim J, Hu C, Moufawad El Achkar C, Black LE, Douville J, Larson A, et al. Patient-Customized Oligonucleotide Therapy for a Rare Genetic Disease. N Engl J Med. 2019 Oct 24;381(17):1644–52.

