## Supplemental Figures for "RNA-sequencing first approach generates new diagnostic candidates in Mendelian disorders"

### Slide 1
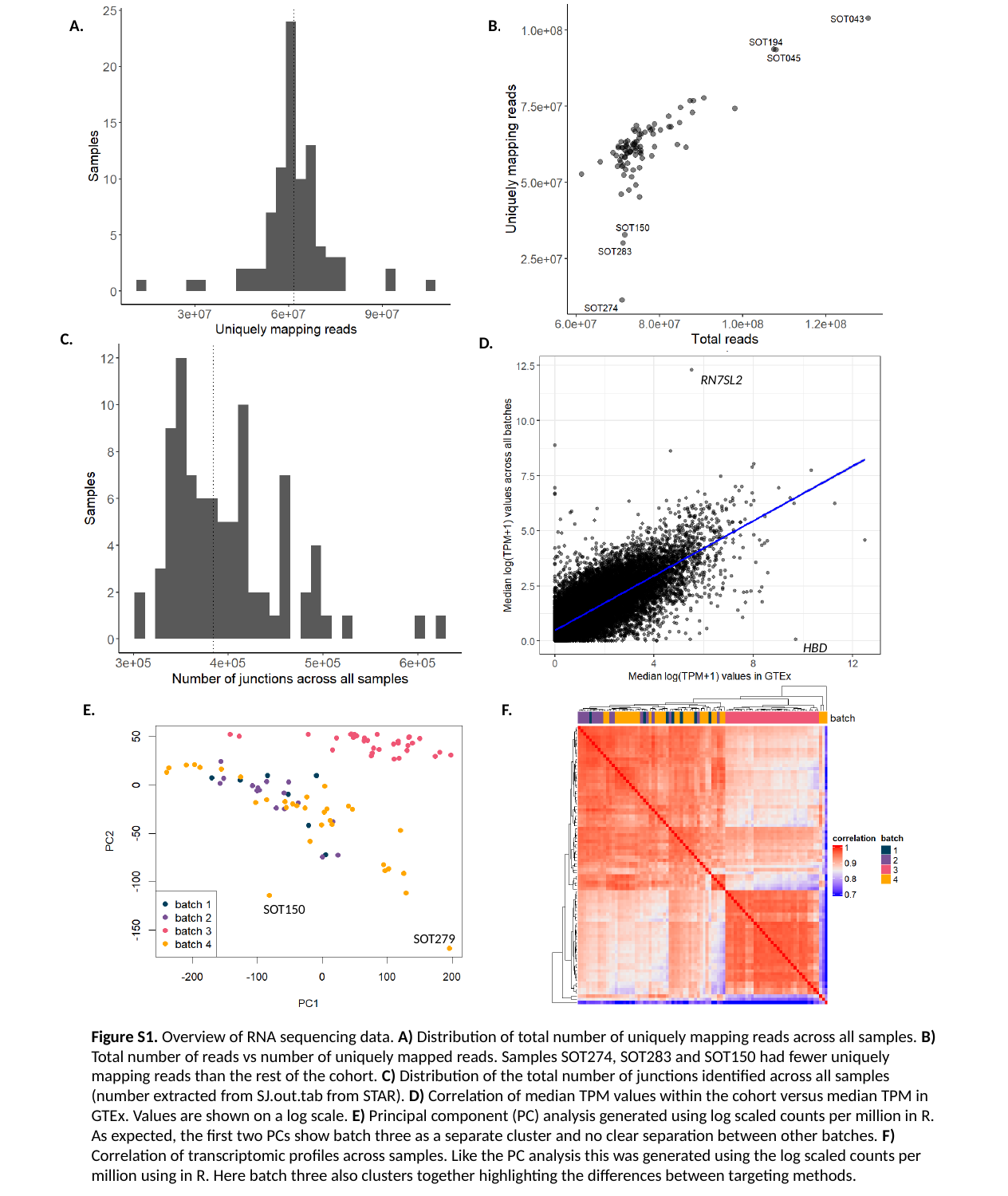

A.
B.
C.
D.
RN7SL2
HBD
E.
F.
SOT150
SOT279
Figure S1. Overview of RNA sequencing data. A) Distribution of total number of uniquely mapping reads across all samples. B) Total number of reads vs number of uniquely mapped reads. Samples SOT274, SOT283 and SOT150 had fewer uniquely mapping reads than the rest of the cohort. C) Distribution of the total number of junctions identified across all samples (number extracted from SJ.out.tab from STAR). D) Correlation of median TPM values within the cohort versus median TPM in GTEx. Values are shown on a log scale. E) Principal component (PC) analysis generated using log scaled counts per million in R. As expected, the first two PCs show batch three as a separate cluster and no clear separation between other batches. F) Correlation of transcriptomic profiles across samples. Like the PC analysis this was generated using the log scaled counts per million using in R. Here batch three also clusters together highlighting the differences between targeting methods.

### Slide 2
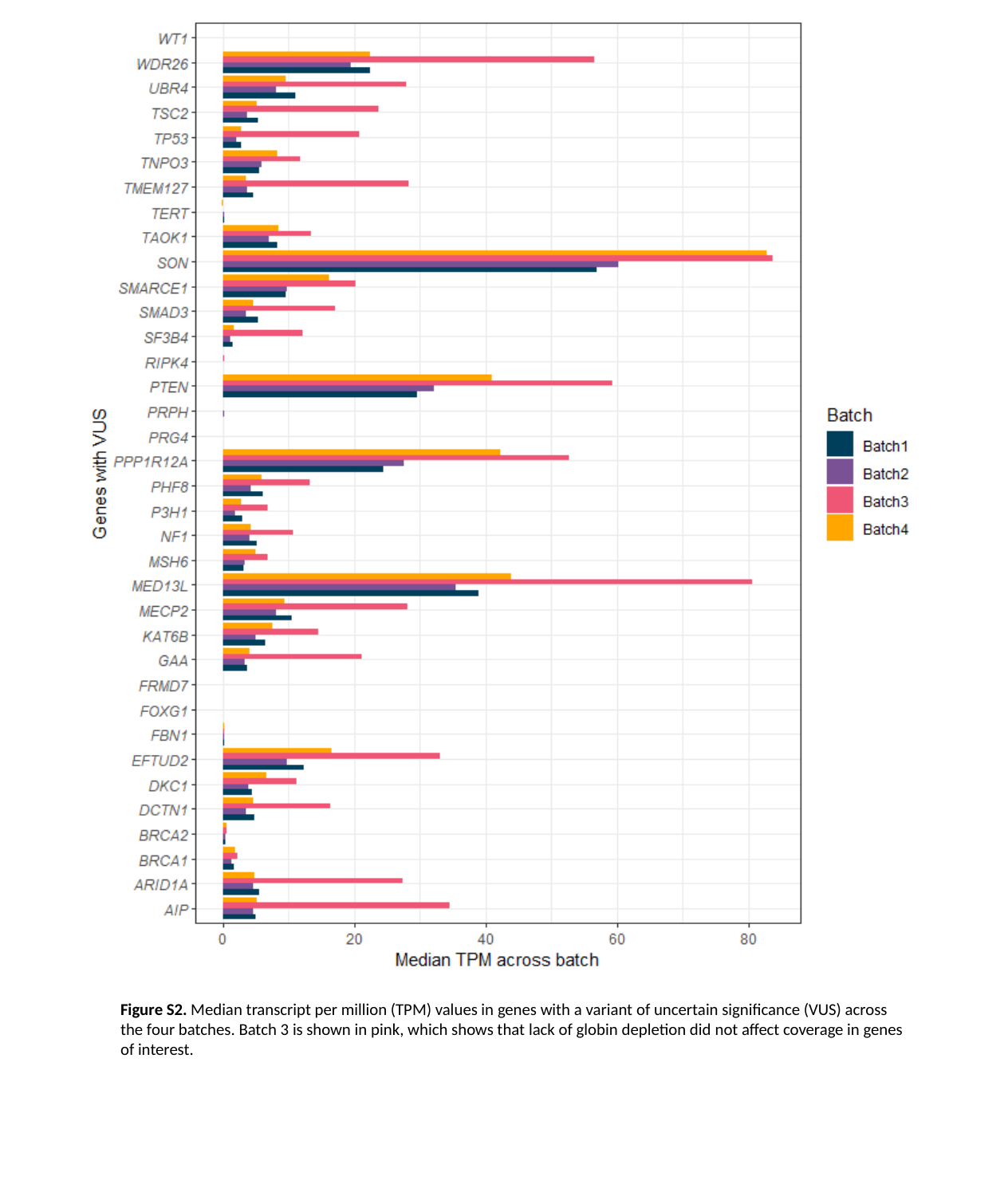

Figure S2. Median transcript per million (TPM) values in genes with a variant of uncertain significance (VUS) across the four batches. Batch 3 is shown in pink, which shows that lack of globin depletion did not affect coverage in genes of interest.

### Slide 3
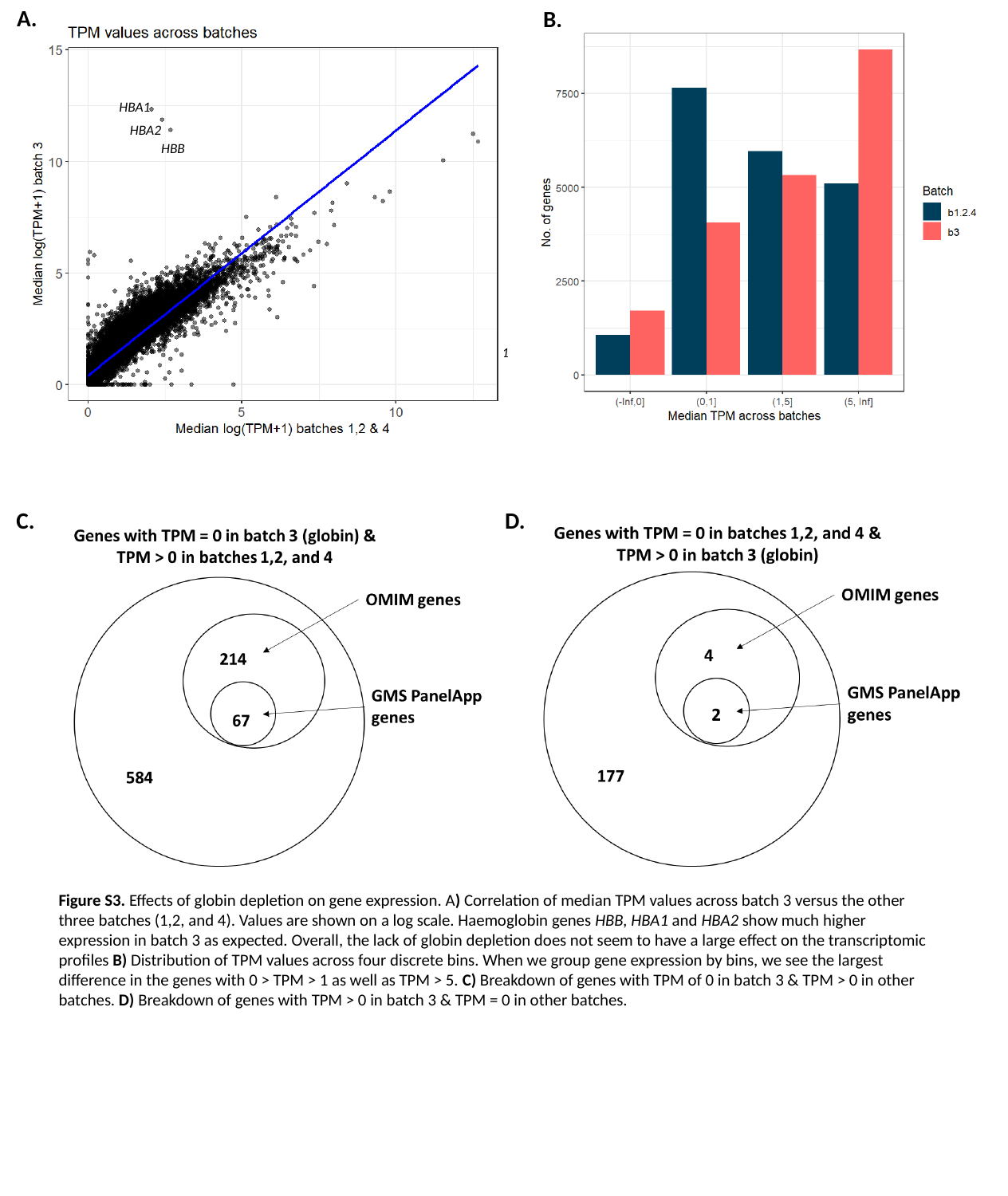

A.
B.
HBA1
HBA2
HBB
HBB
HBA2
HBA1
C.
D.
Figure S3. Effects of globin depletion on gene expression. A) Correlation of median TPM values across batch 3 versus the other three batches (1,2, and 4). Values are shown on a log scale. Haemoglobin genes HBB, HBA1 and HBA2 show much higher expression in batch 3 as expected. Overall, the lack of globin depletion does not seem to have a large effect on the transcriptomic profiles B) Distribution of TPM values across four discrete bins. When we group gene expression by bins, we see the largest difference in the genes with 0 > TPM > 1 as well as TPM > 5. C) Breakdown of genes with TPM of 0 in batch 3 & TPM > 0 in other batches. D) Breakdown of genes with TPM > 0 in batch 3 & TPM = 0 in other batches.

### Slide 4
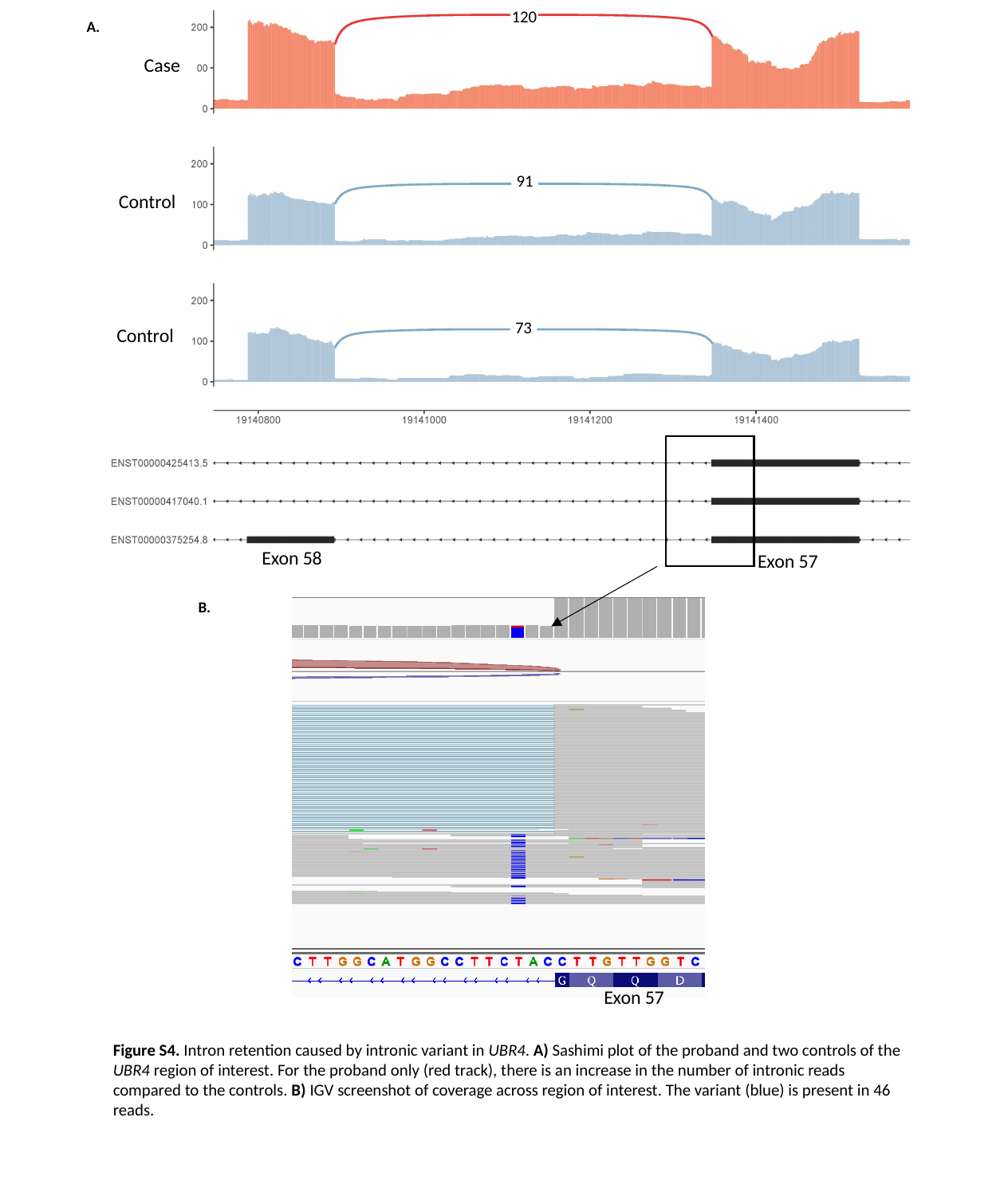

120
A.
Case
91
Control
73
Control
Exon 58
Exon 57
B.
Exon 57
Figure S4. Intron retention caused by intronic variant in UBR4. A) Sashimi plot of the proband and two controls of the UBR4 region of interest. For the proband only (red track), there is an increase in the number of intronic reads compared to the controls. B) IGV screenshot of coverage across region of interest. The variant (blue) is present in 46 reads.

### Slide 5
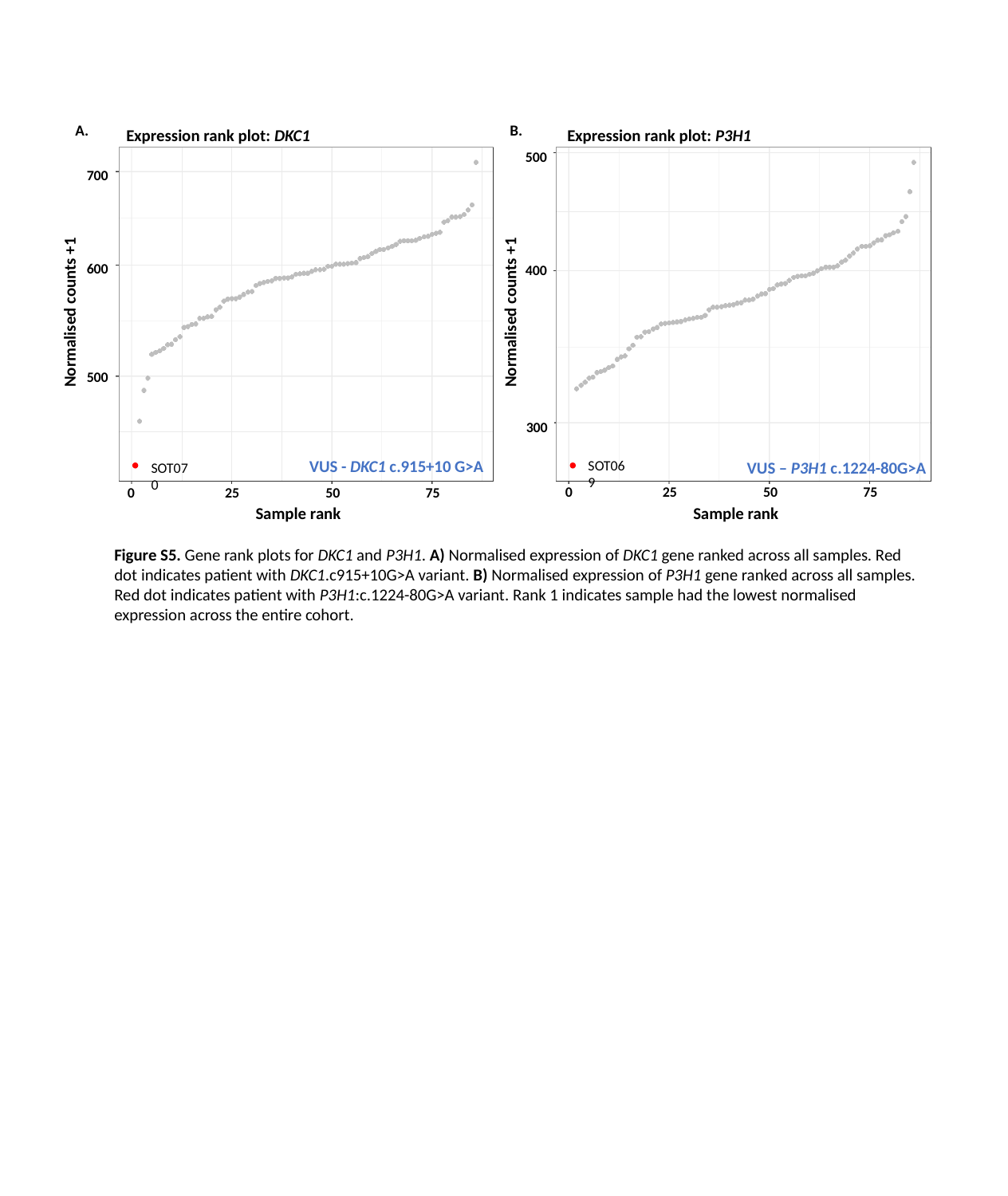

A.
B.
Expression rank plot: P3H1
Expression rank plot: DKC1
500
700
Normalised counts +1
Normalised counts +1
600
400
500
300
VUS - DKC1 c.915+10 G>A
SOT069
VUS – P3H1 c.1224-80G>A
SOT070
0
25
50
75
0
25
50
75
Sample rank
Sample rank
Figure S5. Gene rank plots for DKC1 and P3H1. A) Normalised expression of DKC1 gene ranked across all samples. Red dot indicates patient with DKC1.c915+10G>A variant. B) Normalised expression of P3H1 gene ranked across all samples. Red dot indicates patient with P3H1:c.1224-80G>A variant. Rank 1 indicates sample had the lowest normalised expression across the entire cohort.

### Slide 6
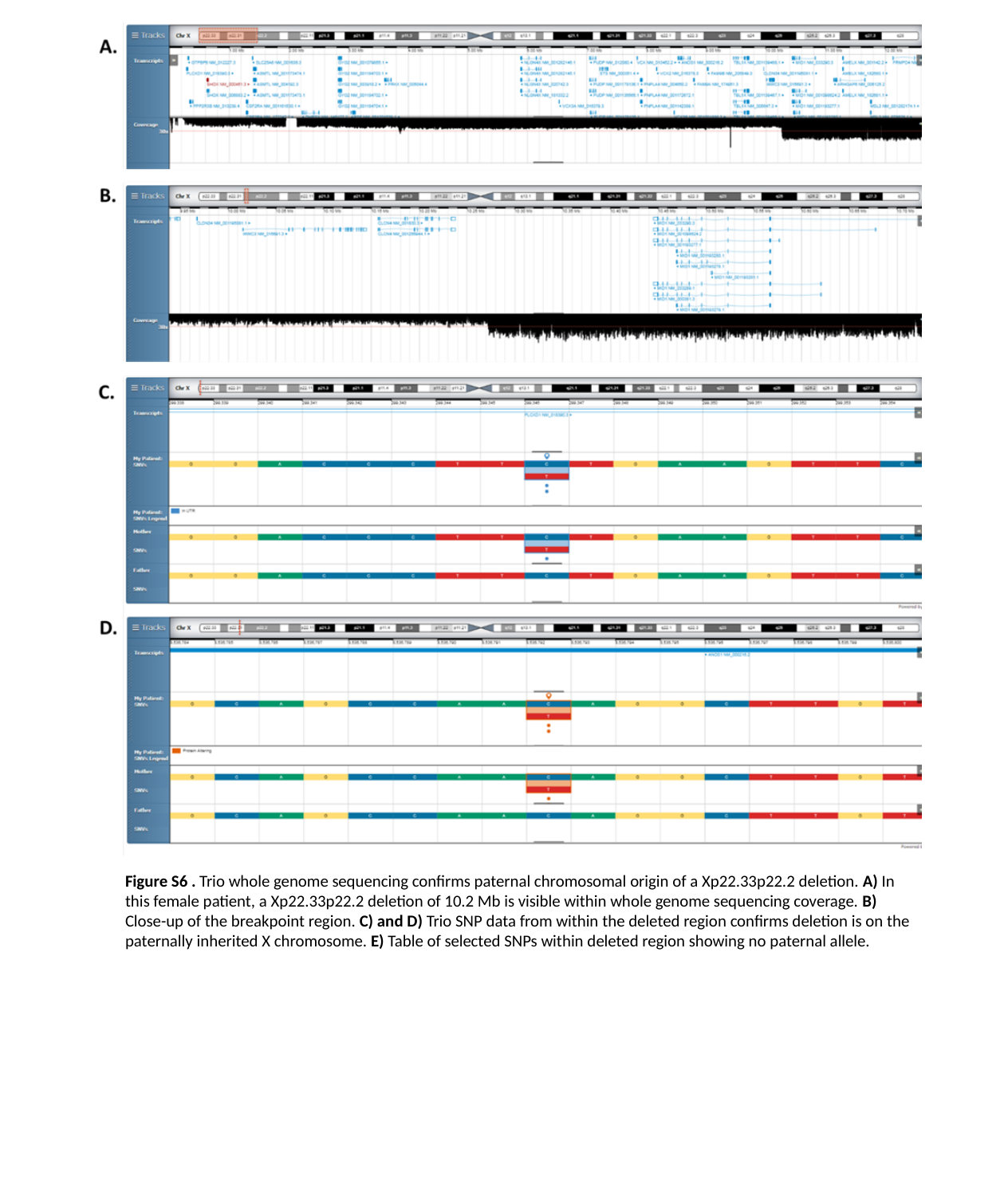

Figure S6 . Trio whole genome sequencing confirms paternal chromosomal origin of a Xp22.33p22.2 deletion. A) In this female patient, a Xp22.33p22.2 deletion of 10.2 Mb is visible within whole genome sequencing coverage. B) Close-up of the breakpoint region. C) and D) Trio SNP data from within the deleted region confirms deletion is on the paternally inherited X chromosome. E) Table of selected SNPs within deleted region showing no paternal allele.
